# Accurate spatial quantification in computational pathology with multiple instance learning

**DOI:** 10.1101/2024.04.25.24306364

**Authors:** Zeyu Gao, Anyu Mao, Yuxing Dong, Jialun Wu, Jiashuai Liu, ChunBao Wang, Kai He, Tieliang Gong, Chen Li, Mireia Crispin-Ortuzar

## Abstract

Spatial quantification is a critical step in most computational pathology tasks, from guiding pathologists to areas of clinical interest to discovering tissue phenotypes behind novel biomarkers. To circumvent the need for manual annotations, modern computational pathology methods have favoured multiple-instance learning approaches that can accurately predict whole-slide image labels, albeit at the expense of losing their spatial awareness. We prove mathematically that a model using instance-level aggregation could achieve superior spatial quantification without compromising on whole-slide image prediction performance. We then introduce a superpatch-based measurable multiple instance learning method, SMMILe, and evaluate it across 6 cancer types, 3 highly diverse classification tasks, and 8 datasets involving 3,850 whole-slide images. We benchmark SMMILe against 9 existing methods, and show that in all cases SMMILe matches or exceeds state-of-the-art whole-slide image classification performance while simultaneously achieving outstanding spatial quantification.

## 1 Introduction

Spatial predictions are critical for computational pathology. For instance, pathologists assess tissue samples visually, and explainable artificial intelligence (AI) tools are expected to produce spatial maps to guide their attention [1–3] The discovery of spatial associations between tissue phenotype and the corresponding genotype can provide biological insights [4, 5], guide biomarker discovery [6, 7], and facilitate downstream tasks such as spatially resolved sequencing [8, 9].

A key bottleneck in the development of spatially-aware computational pathology models is the need for detailed spatial annotations, often unfeasible due to the vast scale of gigapixel images and the need for specialized domain knowledge [10]. Multiple Instance Learning (MIL) has emerged as the leading learning paradigm for whole-slide imaging (WSI) analysis [11], due to its ability to efficiently utilize patient-level labels, which are readily obtainable from pathology reports.

The vast majority of MIL-based computational pathology models adopt a so-called representation-based methodology with an attention mechanism to identify highly discriminative tissue regions, for example for cancer detection and diagnosis [12]. These approaches have been hugely successful, achieving excellent slide-level classification performance. However, attention maps produced by representation-based MIL approaches are often imprecise and unreliable for spatial analysis [13, 14]. This limitation significantly hampers the utility of these methods in scenarios that require accurate phenotypic descriptions beyond a simple global label.

In this manuscript we introduce a new method that performs accurate spatial quantification concurrently with WSI classification, called SMMILe (Superpatch-based Measurable Multiple Instance Learning method). The method is based on *instance*-based MIL equipped with an attention mechanism, a type of MIL with remarkable localization ability [15]. To address the potential limitations of instance-based MIL approaches, specifically their inferior WSI-level prediction capability and misdetection rate, SMMILe uses Neural Image Compression (NIC) [16], two novel instance-based comprehensive attention modules, and a Bayesian refinement network. We investigate the capabilities of SMMILe on 3 diverse families of pathology tasks in WSI analysis, including metastasis detection, subtype prediction, and grading, presented as binary, multi-class, and multi-label classification tasks. We study 8 datasets for 6 cancer types (**Fig. 1a**), including lung, renal, ovarian, breast, gastric, and prostate cancer. We demonstrate that SMMILe achieves state-of-the-art classification performance at the WSI level across cancer types (**Fig. 1b**). Importantly, it simultaneously provides spatial predictions that surpass 9 state-of-the-art computational pathology methods by a large margin (**Fig. 1c**).

**Fig. 1:**
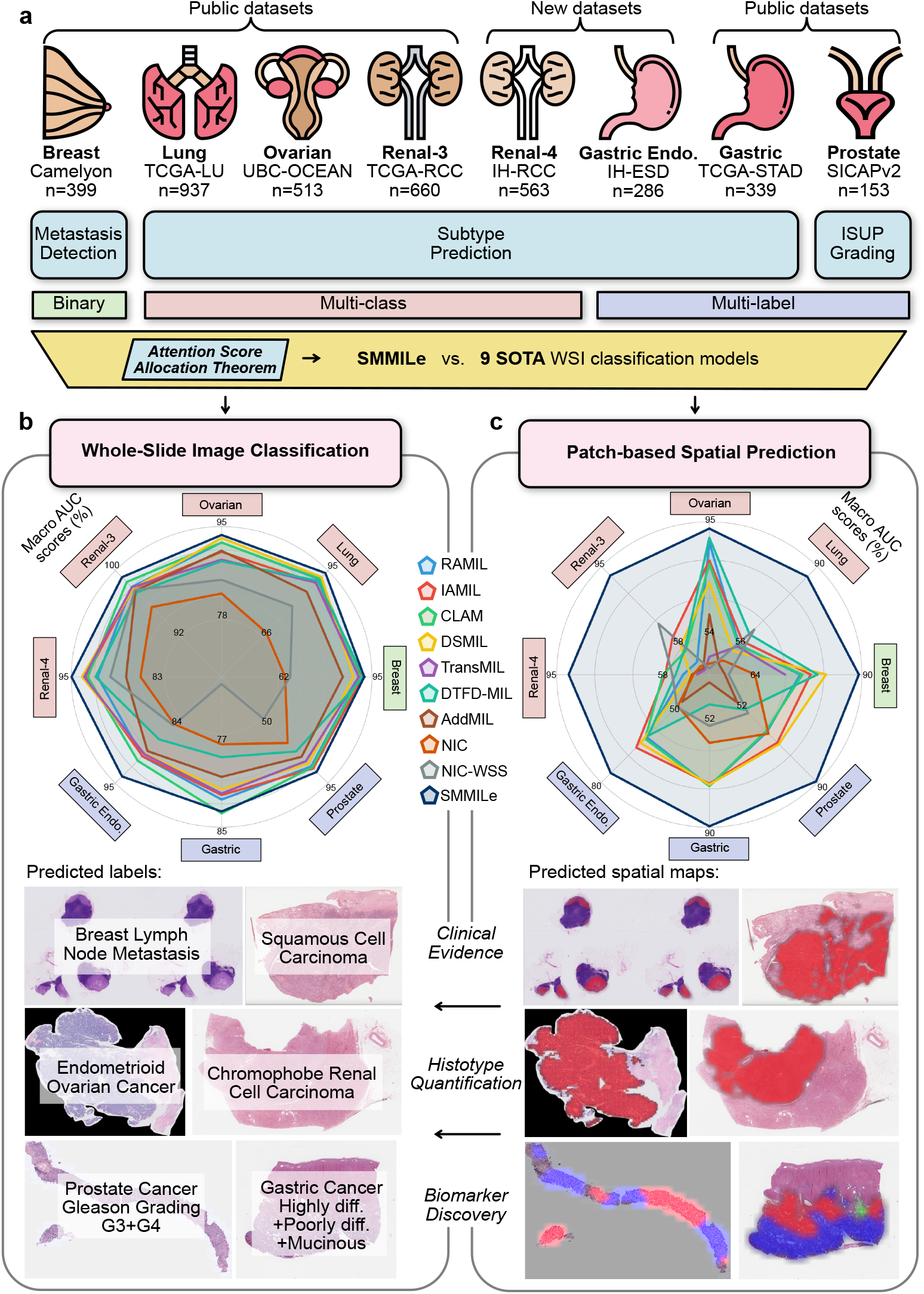
Study Overview and Evaluation. **a**, Study Overview. The distinct attention score allocation mechanisms across different MIL methodologies are theoretically analyzed. Building on this foundation, we present SMMILe, a novel MIL method, and compare it with nine SOTA WSI classification techniques across six public and two in-house datasets. These datasets encompass CPath tasks such as metastasis detection, subtype prediction, and ISUP grading across six distinct cancer types. (see Methods for details on attention score allocation theorem, SMMILe methodology, and evaluation datasets). **b**, Radar plot comparing the WSI-level classification performance of SMMILe and baselines on all evaluation datasets, and examples of WSI predicted labels by SMMILe. **c**, Radar plot comparing the patch-based spatial prediction performance of SMMILe and baselines on all evaluation datasets, and examples of predicted spatial maps by SMMILe. For binary and multi-class classification, red color represents the predicted tumor region; for multi-label classification, different colors represent different predicted phenotypes.

## 2 Results

### 2.1 Instance-based learning produces highly skewed attention maps by design

Despite sharing a foundational MIL framework, instance-based (IAMIL) and representation-based (RAMIL) multiple-instance learning methods diverge in their methodologies for assigning attention scores to individual instances. Understanding these differences is critical to design a framework that has the best of both worlds. With that objective, we analyze the mathematical formulation of the different frameworks by means of three theorems and a targeted synthetic experiment (Section 4.1).

First, we find that, mathematically, RAMIL and IAMIL share a clear mutual link: RAMIL with the most commonly used linear attention is strictly equivalent to Logit-based attention MIL (LAMIL), a variant of IAMIL. The distinction in attention score allocation between LAMIL and IAMIL is closely related to the properties of the output layer’s activation function.

Second, we find that as a consequence of its intrinsic mathematical formulation, IAMIL assigns relatively lower attention scores to low-discriminative and non-discriminative instances, and higher scores to those with highly discriminative power, compared to the allocation patterns observed in LAMIL.

To examine the distinctions between RAMIL and IAMIL in the practical setting, we create a distinguishable synthetic dataset with a ‘positive’ and a ‘negative’ class adhering to the MIL settings (**Fig. 2a, Supplementary Table 1**). Then, we utilize the architecture of AB-MIL [17] as the basis for RAMIL and modify it for IAMIL by conducting attention pooling at the instance level (**Fig. 2b**, see Methods for details of theoretical findings and synthetic experiments). As shown in **Fig. 2c,d**, we find that IAMIL tends to allocate attention scores to highly discriminative instances that are twice higher than RAMIL. However, other positive instances with lower discriminative power have *lower* attention scores in comparison to those in RAMIL. In addition, while the attention scores for most of the *negative* instances in IAMIL fall below 0.003, RAMIL assigns a considerable portion above 0.01 – indicating that the negative instances are assigned much lower attention scores by IAMIL as compared to RAMIL.

**Fig. 2:**
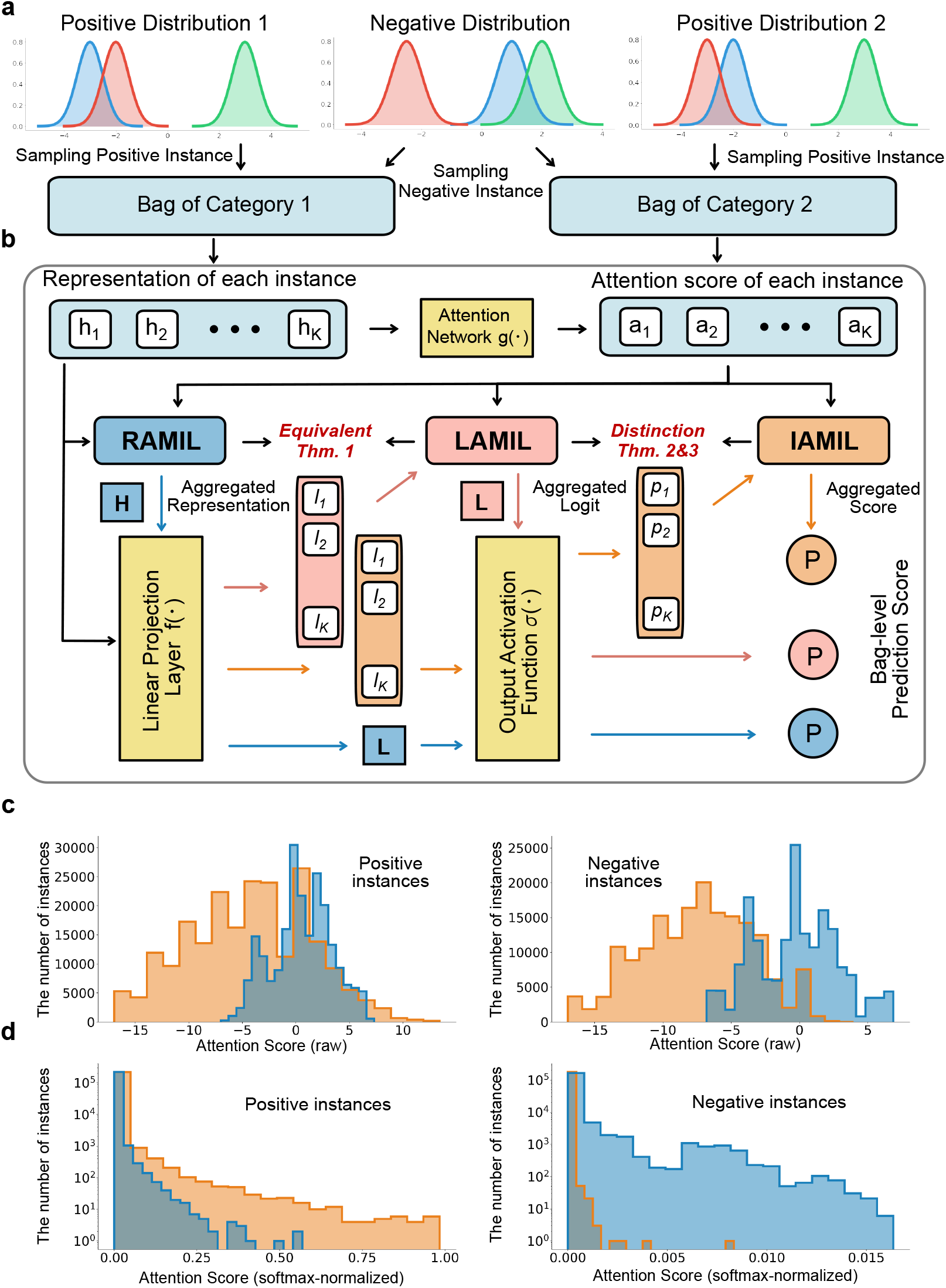
IAMIL versus RAMIL. **a**, Synthetic Data Generation. This synthetic MIL dataset consists of two categories. For these, we designed three types of instance distributions using Gaussian distributions: two are discriminative (positive) and one is non-discriminative (negative). **b**, Frameworks of Three Attention MIL. First, all three methods calculate an attention score for each instance by the attention network. Then, based on these scores, RAMIL combines data at the representation level, LAMIL at the logit level, and IAMIL at the score level. **c**, Histograms of raw attention scores for positive and negative instances. **d**, Histograms of softmax-normalized attention score for positive and negative instances (RAMIL in blue, IAMIL in orange).

Together with our theoretical findings, these observations highlight the key limitation of IAMIL: it produces highly skewed attention maps, focusing only on a limited subset of high-discriminative regions within WSI, leading to a decreased recall rate for the relevant tissue regions. Solving this critical shortcoming is the main barrier for IAMIL to achieve its potential as a superior foundational approach for accurate spatial quantification in WSIs.

### 2.2 Whole slide image classification

Building on from these results, we developed SMMILe, a superpatch-based measurable multiple instance learning method. SMMILe is designed to benefit from the spatial awareness of IAMIL, and equipped with custom modules that address the shortcomings identified previously.

SMMILe comprises a convolutional layer, an instance detector, an instance classifier, and five novel modules (*i*.*e*., the slide pre-processing, the consistency constraint, the parameter-free instance dropout, the delocalised instance sampling, and MRF-based instance refinement). The convolution layer for the instance embeddings enhances their local receptive field. The instance detector, designed with multiple streams, identifies the significance of each instance’s embedding for different categories, facilitating multi-label classification tasks. The classifier assigns each instance’s embedding to its respective category. Ultimately, the bag-level (WSI) classification is determined by aggregating the product of detection and classification scores from all instances (patches).

We evaluated the WSI-level classification performance of SMMILe on eight distinct cancer datasets across three different categories of pathology tasks of increasing complexity.

The simplest category was binary classification. Both instances and bags are categorized into only positive and negative classes (e.g. presence of cancer or not). As an example, we trained the framework to detect breast lymph node metastases using the well-studied Breast (Camelyon16) dataset [18]. We also studied multi-class classification, in which instances and bags can belong to one of a number of positive categories, or to the negative class (e.g. different cancer subtypes vs. no cancer). As an example, we studied four different examples of cancer subtyping tasks: Lung (TCGA-LU, non-small cell lung cancer subtyping) [19, 20], Renal-3 (TCGA-RCC, renal cell carcinoma subtyping, three categories) [21–23], Renal-4 (IH-RCC, renal cell carcinoma subtyping, four categories), and Ovarian (UBC-OCEAN, ovarian cancer subtyping) [24, 25]. Finally, we considered the multi-label classification case, which remains relatively unexplored in previous computational pathology literature. Here a bag may contain instances of multiple positive categories simultaneously, in addition to negative instances (e.g. mixed-type tumour). We evaluated three datasets for heterogeneity tumour analysis: Gastric (TCGA-STAD, gastric cancer subtyping) [26], Gastric Endoscopy (IH-ESD, histotype classification for endoscopic submucosal dissection specimens), and Prostate (SICAPv2, Gleason grading) [27].

All datasets were randomly divided at the patient level into five subsets to facilitate five-fold cross-validation, with the results reporting the mean and variance for each metric.

We benchmarked SMMILe against the two fundamental attention-based MIL methods (RAMIL and IAMIL), in addition to five current state-of-the-art MIL-based WSI classification methods: CLAM [28], DSMIL [29], TransMIL [30], DTFD-MIL [31], and AddMIL [13]. Moreover, we consider two NIC-based WSI classification methods: the standard neural image compression (NIC) approach [16], and NIC-WSS [32], an enhanced variant optimized for regional segmentation. Except for the already tessellated Prostate dataset, we process these WSIs into patches without overlapping. Patch embeddings are extracted from an identical layer (the third residual block) of the ResNet-50, which has been pre-trained on the ImageNet dataset. This approach is uniformly applied across all baselines, including two NIC-based methods initially utilizing an encoder pre-trained on histopathological datasets, paralleling the encoder used in SMMILe to ensure equitable comparison (see **Methods** for details of datasets and implementation).

SMMILe consistently surpasses comparative methods in WSI classification across nearly all datasets for both accuracy and macro AUC score (**Fig. 3a,c**). SMMILe attains accuracy of 87.68%, 94.63%, and 87.32% on the Lung, Renal-3, and Prostate datasets, respectively, exceeding the second-best methods by margins of 3.08%, 2.49%, and 2.92%. In the cases of Breast and Ovarian datasets, while CLAM and DSMIL record the highest accuracy, SMMILe trails narrowly by 1.2% and 0.2% in accuracy, respectively. However, SMMILe secures the top macro AUC scores of 93.21% for the Breast dataset and 90.51% for the Ovarian dataset. The error bars (**Fig. 3a**) and the box plot shapes (**Fig. 3c**) further underscore the robustness of SMMILe in WSI classification across different datasets. The two methods based on NIC demonstrate suboptimal WSI classification performance on most datasets. This shortcoming originates mainly from their intrinsic design, which is intended for handling WSI embeddings generated by pretrained encoders which are finely tuned for discriminative features pertinent to specific domains. These results demonstrate that SMMILe can deliver superior and consistent WSI classification performance across a wide range of cancer types and computational pathology tasks.

**Fig. 3:**
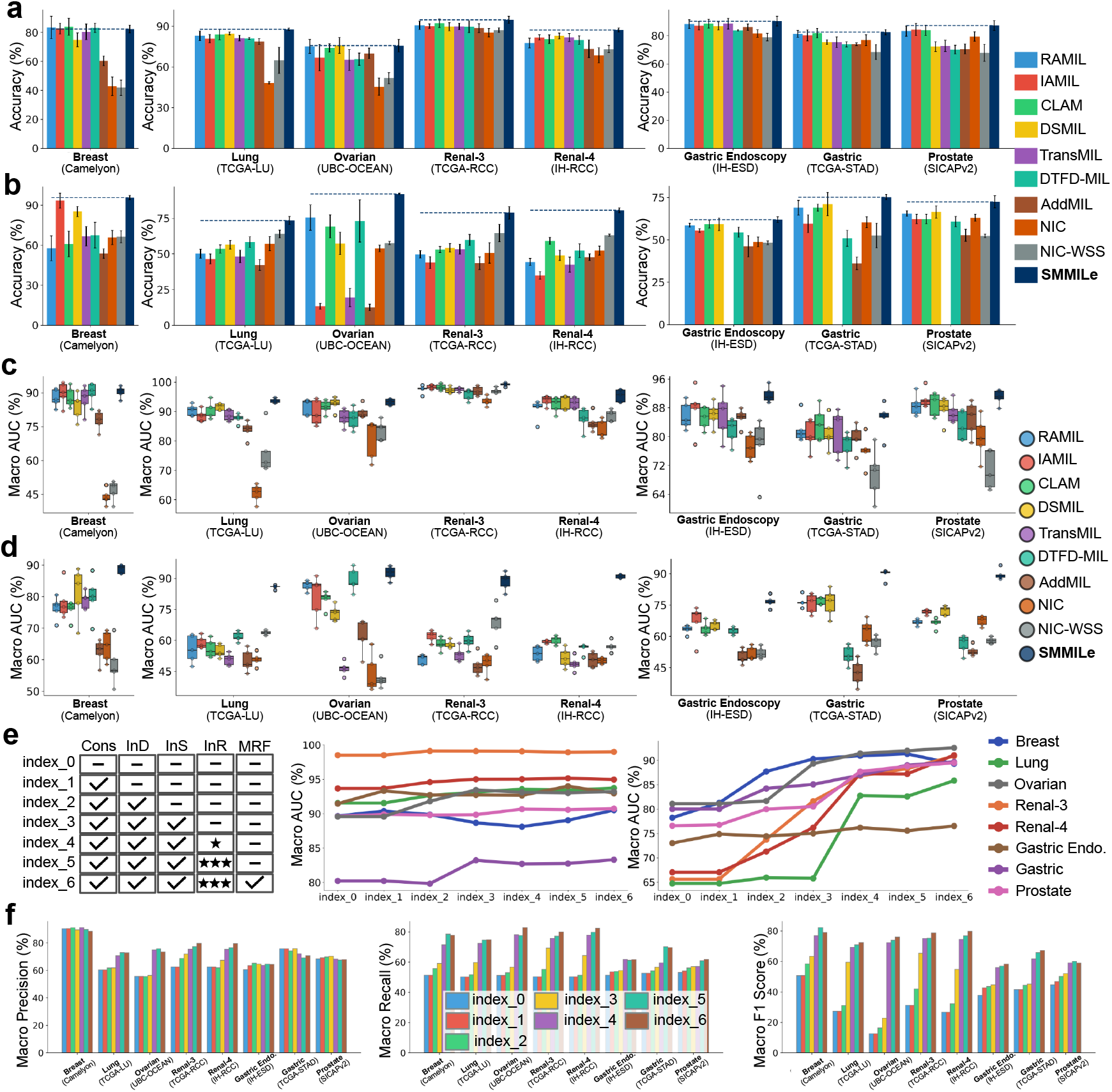
Performance of WSI classification and spatial quantification. **a-d**, the WSI classification (**a**,**c**) and spatial quantification (**b**,**d**) performance of different methods on eight datasets, the accuracy, and macro AUC score are reported. In **a, b**, dashed lines represent the performance of SMMILe. Error bars represent the standard deviation of 5-fold cross-validation. In **c, d**, the individual model performance of each fold is shown via boxplot. **e-f**, the ablation results of SMMILe. From left to right of **e**, the ablation setting of each variation, the macro AUC score of each variation for WSI classification, and spatial quantification, respectively. In **f**, the macro precision, recall, and F1 score of each variation for spatial quantification, respectively.

### 2.3 Spatial quantification and visualization

Beyond the accurate classification of entire WSIs, we wanted to assess the capabilities of SMMILe for spatial quantification, *i*.*e*., patch-level classification, compared to existing methods. Spatial ground truth annotations were available for eight datasets, either in their entirety (Breast, Gastric Endoscopy, and Prostate datasets) or in part (Lung, RCC-3, RCC-4, and Gastric datasets). The Ovarian dataset represents a unique case, with annotations available for only a subset of patches within certain WSIs. Consequently, the evaluation of this dataset is confined to the spatial quantification of these specifically annotated patches. None of the spatial annotations were used for model training in any case.

The derivation of patch-level predictions in representation-based attention MIL baselines (RAMIL, CLAM, DSMIL, TransMIL, and DTFD-MIL) relies on the raw attention scores. In the case of NIC and NIC-WSS, patch-level predictions are acquired from grad-CAM [33] outputs, whereas, for the instance-based MIL methods (IAMIL, and AddMIL), predictions are based on instance scores. SMMILe possesses an instance refinement network capable of directly generating patch-level predictions.

Across all evaluated datasets, SMMILe surpasses other methods by substantial margins (**Fig. 3b,d** and **supplementary tables 3-10**). It achieves macro AUC scores that either exceed or approach 90% in nearly all datasets. An exception is the Gastric Endoscopy dataset, characterized by its extremely imbalanced patient distribution, where SMMILe still demonstrates superior performance. Specifically, SMMILe outperforms the second-best methods by over 20% on the Lung, RCC-3, and RCC-4 datasets; by more than 10% on the Gastric, and Prostate datasets; by nearly 10% on the Breast, and Gastric Endoscopy datasets. In the case of the Ovarian dataset, given the incomplete and highly imbalanced patch-level annotations (a few non-tumor regions are annotated) of this dataset, several methods report commendable accuracy and macro AUC scores. However, while SMMILe only exceeds that of the second-best method (DTFD-MIL) by 3.29% in the macro AUC score, it significantly outperforms DTFD-MIL by 18.94% and 17.09% in the accuracy and macro F1 score, respectively.

Meanwhile, SMMILe exhibits a significant improvement in macro precision, recall, and F1-score (**Supplementary Fig. 4**), on the Lung, Ovarian, Renal-3, Renal-4, and Gastric datasets, because WSIs in these datasets are diagnostic slides of primary tumors, encompassing a vast array of both positive and negative instances. Consequently, SMMILe can effectively mitigate the missed detection and false alarm issues that are prevalent with other baselines.

For the Breast dataset, characterized by a lower proportion of tumor area within each WSI, which means the presence of tumors in only a few positive instances in most WSIs, IAMIL manages to achieve commendable patch-level performance as well. However, the improvement of SMMILe in the Prostate dataset is relatively modest when evaluated in terms of precision, and recall. This dataset, comprising a modest collection of biopsy slides (153 in total), from which only a limited quantity of instances can be generated, does not allow the proposed instance refinement network to be fully leveraged due to the sparse availability of instances for training. Nonetheless, SMMILe has a minimum of 17% improvement in macro AUC scores over other baselines in the Prostate dataset, highlighting its superior capability in distinguishing instances belonging to different histotypes.

Additionally, the spatial visualization results (**Fig. 4**) reveal that both representation-based MIL (RAMIL, CLAM, DSMIL, TransMIL, DTFD-MIL) and NIC-based methods (NIC, NICWSS) incur substantial false positive and false negative errors. Of these, NICWSS demonstrates better consistency with respect to the ground truth. In general, instance-based MIL methods (IAMIL, AddMIL) predominantly identify only a limited number of highly discriminative regions, aligning with our theorem. SMMILe exhibits superior performance in spatial quantification, often producing results nearly indistinguishable from the ground truth, even in challenging multi-label datasets. These results confirm that SMMILe successfully meets both objectives of precise WSI classification and spatial quantification on various datasets.

**Fig. 4:**
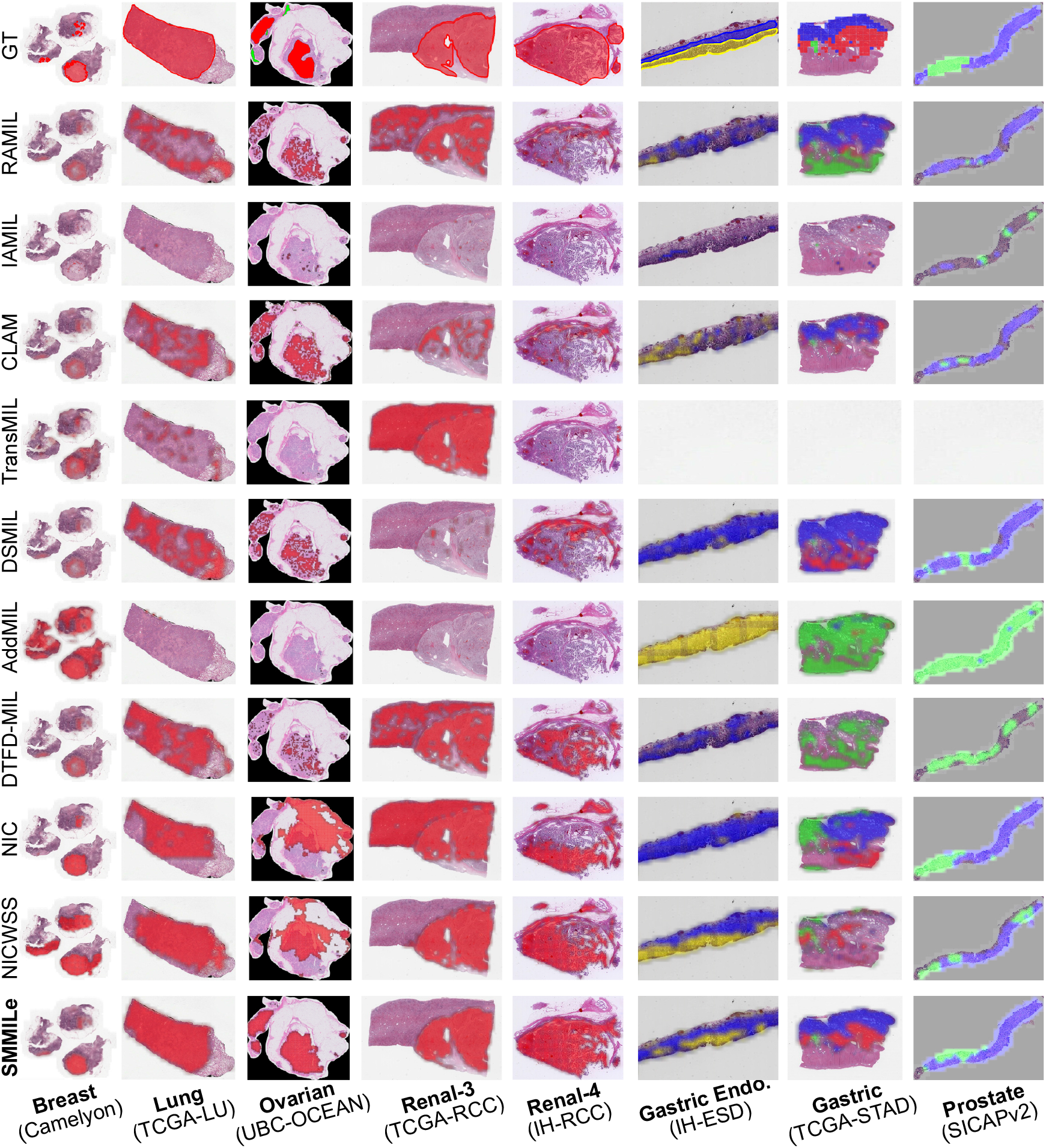
Visualization of spatial quantification across eight datasets. In each dataset, the predicted category of local regions is represented by different colors, whereas regions without color are categorized as normal. The ground truth (GT) delineates the boundaries of tissue belonging to different categories with lines of different colors. For TCGA-STAD and SICAPv2, GTs are presented as patch-level masks. In the UBC-OCEAN case, the green annotation is normal tissue.

### 2.4 Module integration improve SMMILe performance

We conducted comprehensive ablation studies to assess the contribution of each module in SMMILe (**Fig. 5a**), including the consistency constraint, the parameter-free instance dropout (**Fig. 5c**), the delocalised instance sampling (**Fig. 5d**), the instance refinement network (**Fig. 5e**), and the MRF constraint (**Fig. 5e**), each denoted as *Cons, InD, InS, InR*, and *MRF*, respectively.

**Fig. 5:**
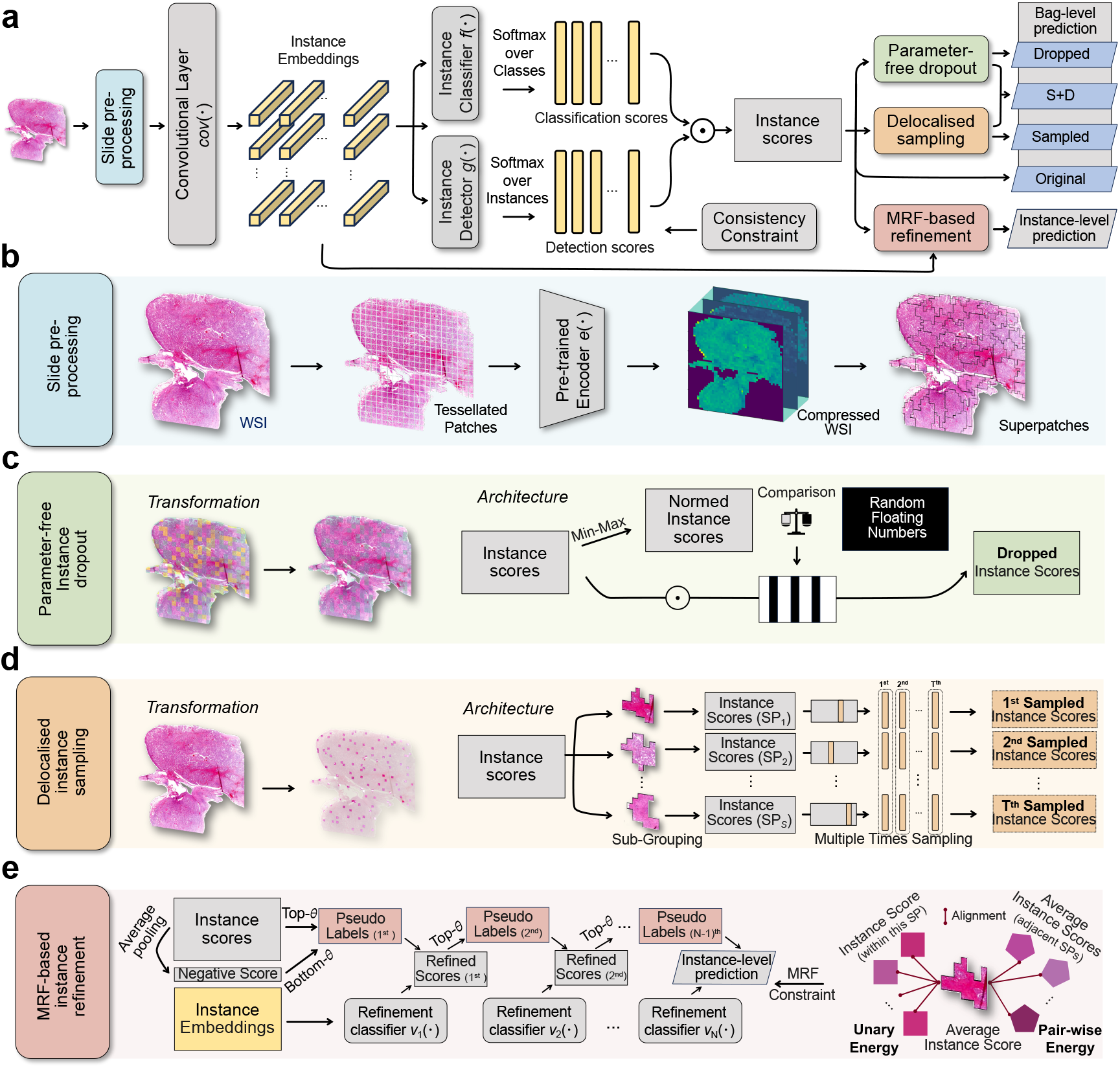
Model Schema of SMMILe. **a**, Overview of SMMILe. **b**, Slide pre-processing. Each WSI is tessellated into patches and mapped into embeddings by the pre-trained encoder. Then, we obtain the compressed WSI based on NIC, which prepares for applying the convolution operation. Concurrently, SMMILe forms a series of superpatches by over-segmentation at compressed WSI. **c**, Parameter-free instance dropout. It involves dropping high-discriminative instances based on their instance scores. **d**, Delocalised instance sampling. It involves generating multiple sub-bags by randomly selecting one instance from each superpatch in multiple rounds. **e**, MRF-based instance refinement. It involves training an auxiliary multi-stage refinement network by self-training strategy with pseudo labels, enhancing the spatial smoothness by MRF energy constraints.

From the WSI classification results (**Fig. 3e**, the first line chart), it is evident that the baseline SMMILe configuration without any modules (index 0) establishes a robust benchmark, attributable to the integration of the NIC feature compression layer with IAMIL. The *InD* (index 2) and *InS* (index 3), acting as two forms of WSI-level augmentation strategies, further enhance the performance of SMMILe across most datasets. However, there is an exception with the Breast dataset, which focuses on breast cancer metastasis detection. Given that many WSIs contain only a few positive instances (*i*.*e*., cancerous patches), *InD* and *InS* might, in certain scenarios, mask all positive instances, adversely impacting the optimization process and the WSI level classification performance of SMMILe. Moreover, the *InR* (index 4 and index 5) and *MRF* (index 6) positively impact WSI classification performance by refining the decision boundaries of the instance classifier, albeit to varying extents. Nonetheless, there are exceptions. The WSI classification in the RCC-3 dataset is relatively straightforward, allowing all methods to achieve high results. Elevating macro AUC performance from 99% to 100% is practically unattainable. Additionally, the sparse presence of positive instances in the WSIs of the Breast and Gastric Endoscopy datasets, along with the limited instance scenario in the Prostate dataset, reduces the effectiveness of *MRF* (index 6). In certain cases, this reduction in effectiveness can even result in decreased WSI classification performance.

From the patch-level spatial quantification results (**Fig. 3e**, the second line chart), it is apparent that every module of SMMILe significantly enhances the spatial quantification performance of SMMILe. Integrating with *Cons* (index 1), which is solely enacted on negative bags, leads to a notable enhancement on the Breast and Gastric Endoscopy datasets, specifically, an increase of 2% and 3% in macro AUC score, respectively. The *InD* (index 2) and *InS* (index 3) modules report enhancements in macro AUC scores ranging from 1-8%, across various datasets. *InR* (index 4 and index 5) exerts a considerable impact across almost all datasets; for example, it facilitates a macro AUC score improvement of approximately 11% for the RCC-4 dataset and 17% for the Lung dataset. Meanwhile, by leveraging spatial smoothness, *MRF* (index 6) contributes to additional improvements in macro AUC scores for all datasets, except for the Breast dataset, where increases of about 1-8% are observed. Furthermore, more detailed metrics regarding spatial quantification (**Fig. 3f**), including macro precision, recall, and F1-score, show that in most datasets the baseline SMMILe (index 0) achieves good precision (**Fig. 3f**, the first bar chart), but the recall is poor (**Fig. 3f**, the second bar chart). This aligns with our theoretical findings, which posit that instance-based attention MIL can accurately identify high-discriminative positive instances, yet often neglects many low-discriminative positive instances. Consequently, it can be noted that each module enhances the recall capability of SMMILe while maintaining high precision, as well as improving the F1 score. It is noteworthy that the *InR* module (index 4, index 5, and index 6), due to its capability to refine and align the decision boundaries for instance classification, contributes the most significant improvement to the performance of SMMILe in terms of macro precision, recall, and F1-score.

## 3 Discussion

To date, relying on patient-level diagnostic labels, which are easy to obtain, a vast number of weakly supervised learning approaches, particularly MIL methods, have been developed for WSI analysis. Most of them utilize attention-based representation-level aggregation for precise WSI classification but overlook the spatial quantification required for many critical tasks and discoveries in clinical pathology. Although these representation-based MIL methods can offer spatial interpretability through the visualization of attention scores, which seems capable of achieving spatial quantification, few works quantitatively assess their interpretability. Here, we conducted a detailed evaluation of WSI-level classification and spatial quantification on 9 SOTA weakly supervised WSI classification methods across 8 diverse datasets across 6 cancer types. In addition to common binary and multi-class classification datasets, we also assessed 3 more complex multi-label classification datasets. We demonstrated that although existing methods can achieve satisfactory WSI classification performance, their spatial quantification capabilities are inadequate for clinical applications and downstream analysis.

In this study, we delved into the theoretical underpinnings of how attention scores are allocated differently across MIL methodologies, specifically between RAMIL and IAMIL. These findings were further corroborated by synthetic experimentation. The theoretical exploration and synthetic experimental evidence suggest that IAMIL, despite its inherent challenges (relatively inferior bag-level prediction capabilities and a high precision yet low recall in positive instance detection) holds superior potential for spatial quantification and interpretability in WSI analysis compared to RAMIL. Subsequently, we illustrated that by integrating NIC, comprehensive attention modules, and an instance refinement network into the IAMIL framework, it is possible to develop a robust MIL method, SMMILe. Compared with other SOTA MIL methods, SMMILe not only excels in superior and consistent WSI classification performance but also provides excellent spatial quantification performance across a wide range of CPath tasks (metastasis detection, subtyping, and grading), with only patient-level labels available for model training.

Furthermore, we demonstrated that the advancements in performance can be directly linked to the innovative modules integrated within SMMILe. Specifically, NIC empowers SMMILe by facilitating convolution operations that broaden the local receptive field of instance embeddings, thereby boosting WSI classification performance. Two advanced comprehensive attention modules (parameter-free instance dropout, and delocalised instance sampling) elevate SMMILe’s ability to recognize positive patches with relatively low-discriminative features, improving its ability to differentiate between such patches and those that are negative. Moreover, given that these two modules incorporate stochastic processes, generating unique instance arrangements for each iteration, akin to WSI-level data augmentation also contributes to enhancing the classification performance and robustness of SMMILe at the WSI level. Building on these enhancements, the instance refinement network addresses a prevalent challenge in attention-based MIL methods: the variability in the optimal decision threshold for attention scores at the patch level across different WSIs. This variability complicates the task of selecting a universal threshold. The network markedly bolsters SMMILe’s ability to discriminate at the patch level by learning a unified decision boundary across different WSIs. When combined with the MRF energy constraint, it further improves the spatial coherence of patch-level predictions, ensuring smoother transitions and consistency in classifications across contiguous patches.

This work has certain limitations. While the evaluation has been performed on eight diverse datasets, it was confined to H&E stained slides for histotype-based classification, due to the available datasets; however, spatial quantification for (1) immunohistochemistry stained slides is also a routine pathological analysis task; and (2) genotype-based classification task could provide profound insights for understanding diseases, while the results of spatial quantification can be validated through spatial transcriptomics. Additionally, the potential of SMMILe has not yet been fully explored. Firstly, the scale of individual datasets is relatively small, with the largest dataset, Lung (TCGA-LU), containing under 1,000 WSIs. Training on larger and more diverse datasets could further enhance its performance. Secondly, the current instance embeddings are solely based on the encoder pre-trained on ImageNet. Combining SMMILe with encoders meticulously fine-tuned on large-scale pathological image databases could further boost its effectiveness.

In conclusion, we believe SMMILe possesses significant potential for clinical application: For general pathology diagnostic tasks, SMMILe not only provides slide-level diagnoses but also offers a comprehensive diagnostic rationale. For tasks involving the quantitative analysis of pathological phenotypes, SMMILe enables precise spatial quantification, further supporting extensive retrospective studies aimed at exploring the relationship between the proportions of different pathological phenotypes and patient treatment responses or prognostic evaluations. For the task of discovering new biomarkers, SMMILe has the potential to identify and quantify previously unknown pathological phenotypes correlated with genotypes, thereby facilitating subsequent targeted and quantitative analyses at the genetic level, such as single-cell and spatial transcriptomics sequencing of subregions.

## 4 Methods

### 4.1 RAMIL versus IAMIL

We delve into the distinct mechanisms of RAMIL and IAMIL, presenting novel theoretical contributions that elucidate their differences. Central to our investigation are two pivotal questions, addressed through three original theorems and a targeted synthetic experiment: (1) How does the variation in the stage of attention aggregation lead to noticeable differences in the attention score allocation? (2) How do these differences contribute to a reduced false positive rate in IAMIL compared to RAMIL?

Our exploration of these questions reveals that, typically in the training process, IAMIL assigns relatively lower attention scores to low-discriminative and non-discriminative instances, and higher scores to those of high-discriminative, in comparison to the allocation patterns observed in RAMIL.

Furthermore, it also disproves some previous interpretations regarding the inaccurate attention results in RAMIL [13]. Contrary to the belief that the attention score allocation in RAMIL has poor interpretability due to instances contributing positively or negatively to bag-level predictions, or due to instance interactions at the classification stage, our findings suggest a different explanation. In RAMIL, bag-level prediction logits can indeed be precisely decomposed into marginal instance contributions as determined by the instance attention scores. However, the properties of the output activation function result in a diminished contribution of these attention scores to the bag-level prediction score. Consequently, during optimization, the loss constraint is limited for attention score allocation, leading to a part of negative instances receiving relatively high attention scores.

#### 4.1.1 Preliminaries

Let {**x**_1_, **x**_2_, …, **x**_*K*_} be a bag of instances (patches) extracted from a WSI **X** ∈ ℝ^*W ×H×*3^, with the WSI-level label **Y** as the supervision, where **x**_*k*_ ∈ ℝ^*D×D×*3^ is a patch represented by its raw pixels and *K* represents the number of instances in this bag. In WSI analysis, representation-based MIL approaches have emerged as the dominant paradigm due to their superior bag-level prediction performance [34–37]. These approaches typically involve three essential steps, which are as follows: (1) Embedding all instances into the same representation space by utilizing the pre-trained encoder *e*(·), a bag of representations is denoted as {**h**_**1**_, **h**_**2**_, …, **h**_**K**_}, where **h**_**k**_ = *e*(**x**_*k*_). (2) Aggregating these instance-level representations into a bag-level representation **H** by a permutation-invariant function, *e*.*g*., mean, and max pooling. (3) Mapping the bag-level representation to prediction score of each category (**P** = {*P*_1_, *P*_2_, …, *P*_*C*_} and *C* stands for the number of categories) using the linear projection function *f* (·) with an output activation function *σ*(·), *e*.*g*., sigmoid, and softmax. Take the mean pooling aggregation as an example, the bag-level prediction score **P** can be represented in the following form:

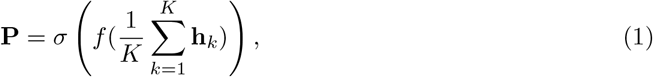

However, representation-based MIL approaches remain a challenge in obtaining individual scores for each instance to ensure interpretability, which is of paramount importance in medical-related applications [38]. An ingenious solution to address this issue is the utilization of attention-based MIL pooling [17], named ABMIL, which incorporates an auxiliary network to generate attention weights for the representation of each instance. This mechanism enables fully trainable and highly flexible MIL pooling, while also providing easily interpretable individual scores for each instance. Consequently, the bag-level prediction score **P** of RAMIL can be reformulated as:

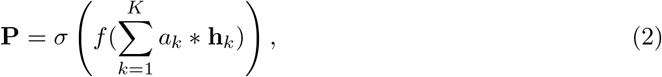

Despite the remarkable achievements of RAMIL, some researchers argued that their visual interpretations are inexact and incomplete [13, 14]. Alternatively, the instance-based MIL approaches adopt the strategy of “mapping first then aggregation”. The bag-level prediction score **P** with the mean pooling aggregation can be formulated as:

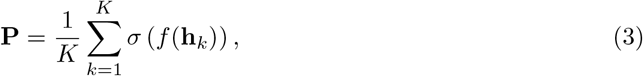

Following this strategy, scores for each instance can be naturally obtained, ensuring high interpretability. Nonetheless, the utilization of instance-based MIL for WSI analysis remains limited. This is attributed to the inaccurate instance labels, *e*.*g*., the pseudo label of non-discriminative instances, during MIL training, which leads to insufficient training of the linear projection function *f* (·) responsible for predicting instance scores in Eq. (3), thereby affecting the bag-level prediction [39]. A straightforward and reasonable solution is to introduce the attention mechanism into the aggregation of instance-based MIL. It differs from RAMIL, where attention scores are generated for the representation of each instance. In this case, the attention mechanism is applied directly to the instance scores. This solution assumes that the attention scores could mitigate the negative impact of inaccurate instance scores. As a result, the bag-level prediction score **P** of IAMIL can be calculated as:

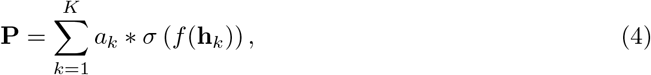

where the logit of *f* (·) for *k*th instance is denoted as *l*_*k*_ = *f* (**h**_*k*_). This formulation is essentially the same as the Multiple Instance Detection Network (MIDN) [15] commonly used in WSOD. MIDN treats the image as a bag and considers the regions generated by object proposal methods as instances. The network is trained with two branches, one for detection and the other for classification, corresponding to *g*(·) and *f* (·) in IAMIL. From Eq. (2) and Eq. (4), the only distinction between RAMIL and IAMIL is identified in the integration stage of the attention mechanism (**Fig. 2b**). Considering observations from substantial existing related works (Section 1), along with the spatial quantification in both methodologies fundamentally rely on the attention scores (*A* = {*a*_1_, *a*_2_, …, *a*_*K*_}), it is reasonable to assume that even this slight distinction between RAMIL and IAMIL can result in notable differences in how attention scores are allocated. This insight prompts us to explore how these two methodologies diverge in their mechanisms for assigning attention scores to individual instances, despite sharing a foundational MIL framework.

#### 4.1.2 Connecting RAMIL with IAMIL

We start by converting the formulation of RAMIL (Eq. 2) into a form that parallels with IAMIL.

##### Theorem 1.

*RAMIL is rigorously equivalent to a specialized variant of IAMIL, wherein aggregation is conducted at the logit level*,

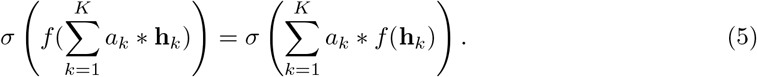

This specific form of MIL termed Logits-based Attention MIL (LAMIL), was initially proposed in [40] (without attention pooling) for WSOD (see **Supplementary Proof 1** for details).

#### 4.1.3 The Gradient Descent Process of Attention MIL

Building upon Theorem 1, the divergence between RAMIL and IAMIL is equivalent to the difference between Eq. (4) and the right side of Eq. (5), *i*.*e*., where the activation function *σ*(·) is applied (Fig. 2). To simplify the following derivation, we extend the RAMIL and IAMIL to more general frameworks. This involves utilizing the class-wise attention mechanism and taking the sigmoid function as the output activation function *σ*(·). Consequently, the attention score *a*_*k*_ is expanded to a vector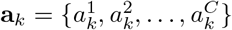, where 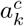 denotes the softmax normalized attention score of the *k*-th instance specific to category *c*. This configuration broadens the applicability of attention MIL across various scenarios, including multi-label classification tasks.

Moreover, since the softmax function leads to entangled relationships between different attention scores across instances, we use the expression of softmax normalized attention scores 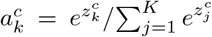 to replace each of them and assess the gradient descent process of 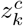 in RAMIL and IAMIL. Let 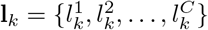 represent the raw outputs (logits) of the linear projection function *f* (·), where **l**_*k*_ = *f* (**h**_*k*_) and each 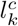 signifies the logits value specific to category *c*. Furthermore, let *L*_*c*_ denote the aggregated logits for category *c*, it can be defined as:

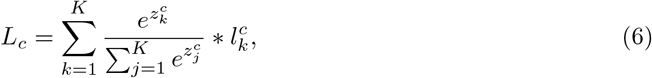

accordingly, the predicted score for category *c* in RAMIL is calculated as follows:

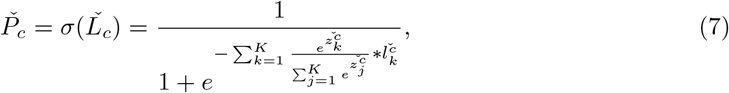

and the prediction score for category *c* in IAMIL is given by:

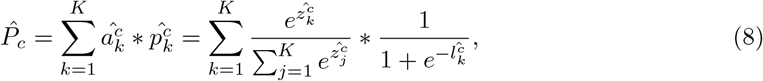

where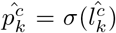. To theoretically explore the variation in the attention score allocation under conditions where RAMIL and IAMIL are optimized to the same level, we posit the following assumptions:

**Assumption 1:** Except for the two attention networks, denoted as *ğ* (·) and *ĝ* (·), both models share identical parameters in their pre-trained encoders *e*(·) and linear projection functions *f* (·), which means 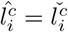 for any instance **x**_*i*_ and category *c*.

**Assumption 2:** Upon training with the same dataset and employing an identical loss function, ∃ *ğ* _*c*_(·), *ĝ* _*c*_(·) with weight 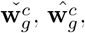 for any *c* ∈ [1, *C*] and *k* ∈ [1, *K* − 1], the following two conditions are satisfied: (1) the bag-level predictions of RAMIL and IAMIL for the same inputs are identical, *i*.*e*., 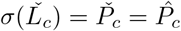, for any category *c*. (2) The gradients of the final losses with respect to the respective network parameters are equivalent,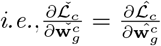.

##### Theorem 2.

*If Assumptions 1 and 2 hold, the relationship in magnitude between* 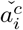 *and* 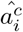 *is directly proportional to the corresponding magnitude relationship between* 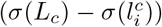 *and* 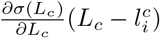, *which is valid for any instance* **x**_*i*_, *i* ∈ {1, 2, …, *K* − 1}:

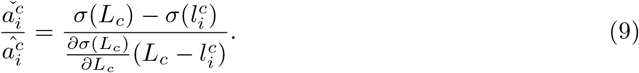

Theorem 2 addresses the first question, asserting that the difference in the attention score allocation between RAMIL and IAMIL is intrinsically linked to the properties of the activation function *σ*(·) across the relevant interval (see **Supplementary Proof 2** for details).

#### 4.1.4 Properties of Activation Function in Local Intervals

Based on Theorem 2, we can infer that if the absolute distance 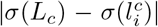 surpasses the projection of 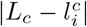 on the tangent line *t*(*l*), then the attention score 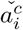 allocated to the *i*-th instance by RAMIL exceeds that assigned by IAMIL 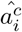, where *t*(*l*) denotes the tangent line to *σ*(*l*) at *L*_*c*_. Conversely, if the projection on the tangent line is greater, the result is reversed.

Also, the activation function (sigmoid) exhibits concavity in the interval [0, ∞) and convexity in the interval (−∞, 0]. This dichotomous property markedly affects the relative magnitudes of 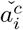 and 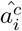. As a result, the magnitude relationship between them is not fixed but is intricately linked to the interval demarcated by *L*_*c*_ and 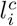.

##### Theorem 3.

*If* 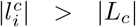*and* 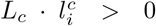, *then* 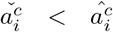.*Conversely, if* 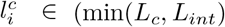, max(*L*_*c*_, *L*_*int*_)), *then*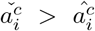. *where L*_*int*_ *is the additional intersection point between t*(*l*) *and σ*(*l*).

Typically, the *i*-th instance is considered to possess high discriminability if 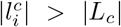 and 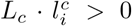. In contrast, the instances are regarded as relatively low-discriminative or non-discriminative instances. Furthermore, owing to the specific properties of the additional intersection point, the conditions outlined in Theorem 3 cover nearly all instances, and few instances are likely to satisfy the rest condition, *i*.*e*., 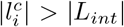 and 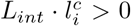 (see **Supplementary Proof 3** for details).

Theorem 3 responds to the second question, highlighting that IAMIL, in contrast to RAMIL, tends to give higher attention scores to high-discriminative instances. Simultaneously, it assigns lower attention scores, which are several times lower, to instances with relatively low discriminability and those non-discriminative instances. Therefore, the instance-level classification results derived from the distribution of attention scores, IAMIL exhibits a very low false positive rate in comparison to RAMIL. However, this also results in only a small part of high-discriminative instances receiving high attention scores, leading to a relatively lower recall rate for positive instances of each category.

#### 4.1.5 The Synthetic Experiment

In the previous section, we assert the near improbability of the condition 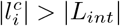 and 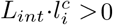. This assertion plays a critical role in delineating the differences in attention score allocation between RAMIL and IAMIL. Consequently, to empirically observe the distributions of *L*_*c*_, and 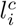 in both RAMIL and IAMIL, as well as to examine the distributions of the raw and softmax normalized attention scores (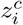, and 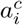), we construct a synthetic dataset that follows the MIL setting (**Fig. 2a, Supplementary Table 1**).

For this synthetic MIL dataset, we generate a set of bags with corresponding bag-level labels where each bag consists of *K* instances, denoted as 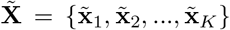. Each instance is a vector with *d* dimensional features, 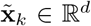. To simplify, this synthetic MIL dataset is defined with two categories. Consequently, we establish three types of instance distributions, used to generate discriminative (positive) instances for two categories, and non-discriminative (negative) instances. Each instance distribution is comprised of Gaussian distributions with different means and variances for each of the *d* dimensions, from which we sample each feature of instances. Therefore, the bag of each category is composed of a random proportion *r* of positive instances belonging to that category, supplemented with the remaining negative instances. It is important to note that the positive instance distributions for the two categories are significantly different, while their distributions, compared to the negative instances, are similar but non-overlapping. This setup is designed to synthesize positive instances with high and low discriminative features. Ultimately, the dataset comprises a total of 2,00 bags, each containing 1,000 instances. The ratio of positive instances within each bag is randomly sampled from a range of 0.1 to 1. The dataset is evenly distributed across the two categories, with each category consisting of 100 bags.

For model training, we utilize the architecture of AB-MIL[17] as the basis for RAMIL and modify it for IAMIL by conducting attention pooling at the instance level. Given that the instance 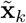 is not an image, we omit the embedding step. Only the linear projection function *f* (·), with 4 hidden nodes, and the gated attention network *g*(·), with 8 hidden nodes in the first layer and 4 hidden nodes in the second layer, are used and trained. The loss function employed is binary cross entropy computed for each category. The Adam optimizer is used, with the learning rate set to 0.001. To ensure a consistent level of convergence between RAMIL and IAMIL, the training process is stopped when the loss decreases below a fixed threshold, specifically 0.05.

We can observe that when the model converges, the values of |*L*_*c*_| for most bags are greater than 5 (**Supplementary Fig. 3(a)**), resulting in corresponding |*L*_*int*_| values surpassing 144 (**Supplementary Fig. 2(c)**). The values of 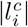 for most negative instances are less than 5 (**Supplementary Fig. 3(b)**), which means they are positioned within the interval demarcated by *L*_*c*_ and *L*_*int*_. Also, the values of 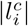 for a few positive instances are greater than |*L*_*c*_| (**Supplementary Fig. 3(c)**). Following Theorem 3, this indicates that IAMIL assigns lower attention scores to the majority of negative instances, while a minority of positive instances receive higher attention scores compared to those assigned by RAMIL. This distinction is also evident from the distribution differences of 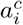 and 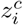 in RAMIL and IAMIL (**Supplementary Fig. 3(d-g)**).

### 4.2 SMMILe

Reiterating the definition in Section 4.1, a set of patches {**x**_1_, **x**_2_, …, **x**_*K*_}, each of size *D* × *D*, are extracted from a WSI **X** of size *W* × *H*, with the corresponding label *Y* . The objective of SMMILe is to learn a transformation function that maps the set of patches {**x**_1_, **x**_2_, …, **x**_*K*_} to the WSI-level label **Y**. Concurrently, it also aims to predict instance-level labels *y*_1_, *y*_2_, …, *y*_*K*_ for each individual patch.

In MIL settings, supervision is exclusively available at the WSI level. Previous research has predominantly focused on binary or multi-class classification, where each WSI is assigned to a single category, *i*.*e*., **Y** is represented as a scalar in binary classification or as a *C*-dimensional onehot vector in multi-class classification. In this paper, we expand SMMILe to accommodate multilabel classification. Consequently, **Y** = {*Y*_1_, *Y*_2_, …, *Y*_*C*_} is configured as a *C*-dimensional vector, with each element *Y*_*c*_ being a binary indicator that is independently distributed, representing the presence or absence of each category in this WSI. This adaption aligns SMMILe with more general CPath scenarios, capturing multiple phenotypic categories that may concurrently exist in a single WSI.

#### 4.2.1 Network Architecture

The proposed network architecture (**Fig. 5**) begins with a pre-trained encoder *e*(·), mapping all instances {**x**_1_, **x**_2_, …, **x**_*K*_} to a uniform embedding space {**h**_1_, **h**_2_, …, **h**_*K*_}. Given the large number of instances contained within each WSI, the encoder training becomes time-consuming and computationally strenuous, leading to the parameters of the encoder generally being kept frozen. In this paper, we employ the ResNet-50 [41], pre-trained on ImageNet [42], as our encoder *e*(·). Feature maps are extracted following the third residual block and aggregated through global average pooling, producing a 1024-dimensional embedding for each instance. This pre-trained encoder has been demonstrated to be effective in numerous RAMIL studies for CPath [28, 30, 31, 43].

Subsequently, a convolutional layer *cov*(·) is introduced, which further maps the instance embeddings to a lower dimension. The parameters of *cov*(·) are trainable for increasing the flexibility of the entire MIL framework. While existing works often employ a linear projection layer for this function, we replace it with a convolutional layer to enhance the representation ability of our framework for downstream tasks by introducing WSI-level local receptive fields. Similar to NIC [16], we reposition the instance embeddings {**h**_1_, **h**_2_, …, **h**_*K*_} according to their positions in the WSI, and fill other positions with zero embeddings, creating a compressed WSI 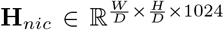 to enable the convolution operation. We utilize 128 convolutional kernels with size 3 × 3 and padding operation, ensuring that the size of the compressed WSI is maintained after convolution, *i*.*e*., 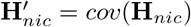, where 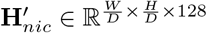. Then, the compressed embeddings 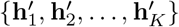 of all instances can be obtained from 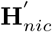 based on their respective positions. It is important to note that when the size of convolutional kernels is set to 1 × 1, *cov*(·) degrades to a linear projection layer, which is particularly effective for specific MIL tasks with very limited positive instances in each bag, such as cancer metastasis detection.

In the final stage, the compressed embedding of each instance 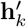, serves as the input for an instance detector *g*(·) and an instance classifier *f* (·). The instance detector, analogous to the attention network in RAMIL frameworks, utilizes the gated attention mechanism [17]. It comprises three linear projection layers with 64, 64, and *C* hidden nodes, respectively, assigning category-wised raw attention scores 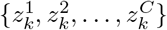 to each instance. Then these raw attention scores 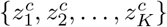 of each category are normalized via softmax function across all instances, resulting in detection (attention) scores 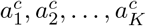, where the sum of them remains invariant to *K* and equals to 1. The instance classifier *f* (·) is a linear projection layer with *C* hidden nodes. It maps the compressed embedding 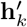 of each instance to category-related scalars 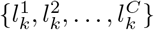. These scalars are then normalized over categories using the softmax function for multi-class classification, or the sigmoid function for binary and multi-label classification, yielding the classification scores 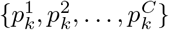 for each instance.

The bag-level prediction score *P*_*c*_ for each category is then obtained by taking the dot product of classification and detection scores, and summing them up, as defined in the aggregation formula in Eq. (4).

#### 4.2.2 Instance-based Comprehensive Attention

To enhance the comprehensive attention capability of SMMILe toward all discriminative instances, we adhere to the traditional MIL by categorizing bags into two types: negative bags and positive bags. Negative bags do not contain any positive instances of any category, as exemplified by WSI of normal tissue; whereas positive bags include positive instances of one or several categories. We propose (1) an attention consistency constraint for negative bags; (2) a parameter-free instance dropout module; and (3) a superpatches-based delocalised instance sampling module for positive bags. It is worth noting that the instance dropout and instance sampling modules presented in this section introduce diversity in instance combinations of each bag, effectively serving as two forms of bag-level augmentation. This, in turn, enhances the performance of bag-level predictions.

##### Consistency Constraint

A bag is classified as negative only if none of the instances within it belong to a positive category. Thus, the classification of negative bags should not rely on a subset of instances but rather on all instances. Instead of applying the same attention mechanism to both negative and positive bags like previous RAMIL approaches, we introduce a consistency constraint for the attention mechanism, which restricts all instances in a negative bag should having the same attention score. This consistency loss is defined as follows:

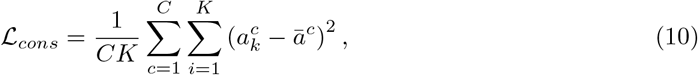

where 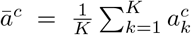. By applying this MSE loss penalty, the attention scores across all instances in a negative bag become uniform, effectively ensuring that no individual instance makes special contributions to any category. This uniformity explicitly boosts the classification accuracy for negative bags as well as the recognition ability of SMMILe to negative instances, thereby implicitly improving the comprehensive attention for discriminative instances.

##### Parameter-Free Instance Dropout

The comprehensive attention capability of SMMILe is limited by focusing mainly on high-discriminative instances and neglecting others. An intuitive idea is that during the training process, omitting high-discriminative instances while retaining bag-level supervision could encourage the model to focus on the remaining discriminative instances. This approach introduces an additional challenge: determining the ideal instance dropout rate. Specifically, excessive instance dropout during early, unstable training phases may obstruct model convergence. In contrast, a low dropout rate in stable phases offers limited benefits. Additionally, with substantial variation in positive instance proportions across different bags, selecting a uniform dropout rate effective for all bags is unfeasible.

To resolve this issue, we design a parameter-free instance dropout module (**Fig. 5c**) Instead of applying dropout to attention scores, which do not directly reflect the contribution of each instance towards the bag-level prediction score, this module targets the instance scores (*i*.*e*., the product of classification and detection scores for each instance), which have strict marginal contributions to the bag-level prediction score [13]. For a set of instance scores 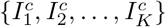 of a bag, where 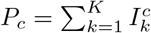, and 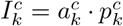, we first apply Min-Max normalization to obtain normalized instance scores 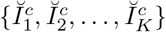. Then, a corresponding set of random floating-point numbers 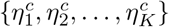, *η* ∈ [0, 1] is generated. We compare them pairwise to obtain the instance drop masks 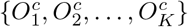. Finally, these instance drop masks are applied to the instance scores, and the bag-level prediction score with instance dropout for category *c* is computed as:

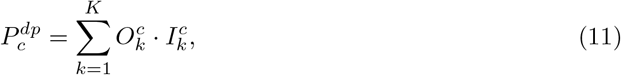

where 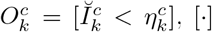 denoting an Iverson bracket. It can be observed that the proposed instance dropout module does not require any additional hyperparameters. The decision to drop an instance is based on its instance score 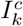. The higher 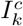, the less likely it is to meet the condition 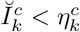, making it more prone to be dropped.

##### Superpatch-based Delocalised Instance Sampling

Instance sampling emerges as another viable solution for enhancing the comprehensive attention capability of SMMILe. By performing multiple rounds of random sampling within a bag, each sampling generates a pseudo-bag composed of a subset of instances. The bag-level supervision is then applied to guide predictions for these pseudo-bags. This approach enables the model to focus on diverse sets of discriminative instances in each pseudo-bag, thus improving its comprehensive attention ability. Nevertheless, this kind of random sampling is uncontrollable and may lead to some pseudo-bags lacking positive instances, thereby introducing substantial noise into the training process of the model. Here, we propose a superpatch-based delocalised instance sampling module (**Fig. 5d**) to address this issue. Recall the compressed WSI **H**_*nic*_ we constructed for a bag, wherein each pixel corresponds to a patch in the original WSI. We employ the Simple Linear Iterative Clustering (SLIC) [44], a widely-utilized, non-trainable clustering-based partition algorithm, to generate a set of superpatches {SP_1_, SP_2_, …, SP_*S*_} from **H**_*nic*_, where *S* indicates the number of superpatches of each bag ((**Fig. 5b**)). This leads to patches that are spatially close with similar representations being grouped into the same superpatch. Based on these superpatches, instances of a bag can be divided into *S* subsets. By conducting *T* rounds of random sampling with replacement, where each round involves sampling one instance from each subset to create a pseudo-bag with *S* delocalised instances, a total of *T* pseudo-bags are generated. The instance sampling is also performed on instance scores directly. For *t*-th pseudo-bag, we have the sampled instance scores 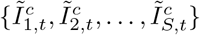 for category *c*, where 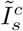 is an instance score sampled from superpatch SP_*s*_, and the bag-level prediction score of *t*-th pseudo-bag is calculated as:

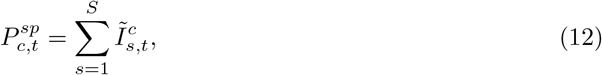

Owing to the characteristic of superpatch, each pseudo-bag is composed of instances exhibiting varied spatial and representation distributions, providing the necessary diversity to encompass both positive and negative instances. Furthermore, sampling instances randomly from superpatches in each round results in a diverse array of instance combinations. This variety mitigates the overshadowing effect of high-discriminative instances on low-discriminative instances within the same pseudo-bag, consequently encouraging the model to focus more on those low-discriminative instances. Additionally, integrating instance sampling with instance dropout is particularly beneficial in scenarios where positive instances are predominant within a bag, such as subtyping on primary tumor slides. The bag-level prediction score for the *t*-th pseudo-bag, when instance dropout is applied, is computed as follows:

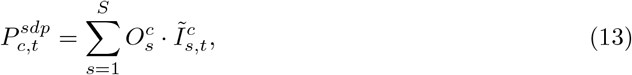

where 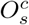 is the instance drop mask for 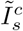. This can be regarded as masking high-discriminative superpatches.

Finally, all bag-level predictions generated by SMMILe, *i*.*e*., 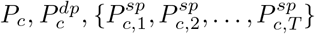, and 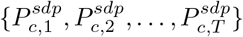, are supervised by bag-level label *Y*_*c*_ of each category. The classification loss for each bag is calculated as:

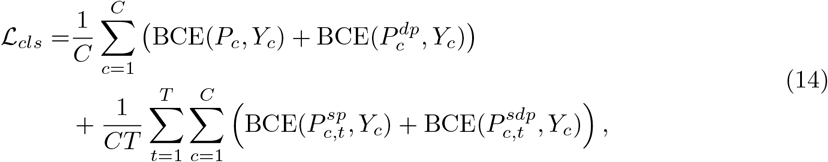

where BCE(*P, Y*) stands for the binary cross entropy loss between the prediction *P* and the true label *Y* .

#### 4.2.3 MRF-based Instance Refinement

The aforementioned modules endow SMMILe with the capability to distinguish between positive and negative instances within each bag for different categories. However, due to the diversity among different bags, such as variations in feature distributions and significant differences in the proportions of positive instances, it is nearly impossible to choose a unified decision boundary for instance-level classification across varying bags and categories. Consequently, we design an instance refinement network to align features of instances of the same category within different bags, thereby enabling us to learn a unified instance-level classification decision boundary across diverse bags.

##### Instance Refinement Network

The instance refinement network is structured with *N* linear layers {*v*_1_(·), *v*_2_(·), …, *v*_*N*_ (·)}. Each layer is implemented with (*C* + 1) hidden nodes and employs a softmax activation function for output generation. Here, *C* represents the number of categories for WSIs, excluding the negative category. For the negative WSI, the label **Y** is encoded using a *C*-dimensional vector of zeros. Consequently, the term (*C* +1) incorporates the negative category to account for instances. These linear layers are assigned the identical task of generating predictions for individual instances but involve different sets of instances with associated compressed embeddings and pseudo-labels for training.

The challenge lies in acquiring pseudo-labels for network training. Leveraging the distinguishing capability of SMMILe for instances of different categories within each bag, we propose an online sample selection and labeling strategy. During each epoch of training, we can obtain a set of instance scores 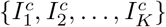 for category *c* from SMMILe. From this set, we select the top *θ* percent of instances, labeling them as belonging to category *c*. This selection is restricted to the categories present in each bag. For negative samples, we first compute the mean score across different categories for each instance, represented as 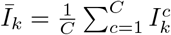. Subsequently, we select the bottom *θ* percent of instances from these averages {*Ī*_1_, *Ī*_2_, …, *Ī*_*K*_ }, labeling them as negative, *i*.*e*., category (*C* + 1). Consequently, the first linear layer *v*_1_(·) is trained using the compressed embeddings and pseudo-labels of the selected instances, represented as 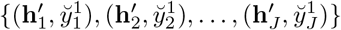, where *J* denotes the total number of selected instances, and 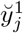 is the first-round pseudo-label of *j*-th selected instance.

Building on the concept of self-training, we employ the prediction results of instances from *v*_1_(·) to supervise the learning of *v*_2_(·), and similarly for subsequent layers, to achieve a higher degree of instance refinement. Specifically, the compressed embeddings of all instances in a bag are fed into *v*_*n*_(·), yielding prediction scores of these instances, denoted by 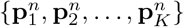, where 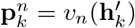is a (*C* +1)-dimensional vector, where each element represents the probability of the instance belonging to a corresponding category. Then, the proposed online sample selection and labeling strategy is employed here, the selected instances and (*n*+1)-round pseudo-labels are used to train the subsequent linear layer *v*_(*n*+1)_(·). It is crucial to note that since each linear layer in the instance refinement network can generate predictions for the negative category (*C* + 1), there is no requirement for a separate selection process for negative samples, except for the training process of *v*_1_(·). Also, the instances selected for each linear layer are likely to differ, while the total number *J* remains constant. Through this strategy, every linear layer in the instance refinement network can be concurrently trained with the SMMILe primary network in each epoch. The refinement loss is defined as:

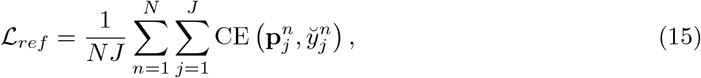

where CE(**p**, *y*) stands for the categorical cross entropy loss between the prediction **p** and the pseudo-label *y*. The optimization process of the instance refinement network also facilitates learning more uniform discriminative instance features (generated by *cov*(·)) across different bags, which enhances the bag-level prediction performance of SMMILe.

##### superpatch-based MRF Constraint

Nevertheless, the proposed sample selection strategy, which treats each instance as an independent entity and only high-scoring instances from each category can be used for supervision, may induce prediction biases in the instance refinement network. Moreover, the current instance refinement network ignores the spatial relationship between instances (patches) within a bag (WSI), which is important for the comprehensive detection of positive instances. Therefore, we introduce a superpatch-based MRF constraint that incorporates local spatial smoothness at the WSI level. This constraint requires the minimization of both the first-order energy within each superpatch and the second-order energy between adjacent superpatches. Consider the *n*-th linear layer *v*_*n*_(·). The prediction scores for instances within the superpatch SP_*s*_ are represented as 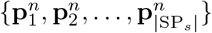, where |SP_*s*_| signifies the count of instances in this superpatch. Furthermore, the prediction score for superpatch SP_*s*_ is calculated as 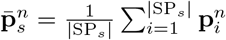, and it is surrounded by *M*_*s*_ adjacent superpatches, whose prediction scores are denoted by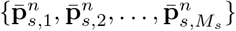. The MRF constraint loss for superpatch SP_*s*_, incorporating both first-order and second-order energy, is defined as follows:

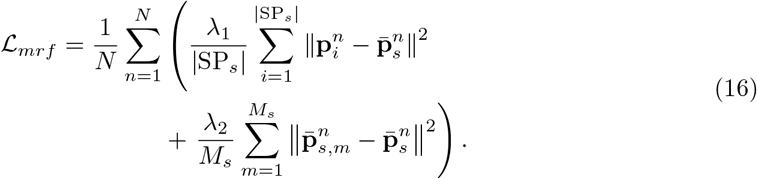

where *λ*_1_ and *λ*_2_ control the balance between first-order and second-order energy. This constraint can implicitly propagate the pseudo-label information of high-scoring instances to the local regions constrained by superpatches, thereby enhancing the spatial smoothness of instance prediction scores.

### 4.3 Evaluation Datasets

**Breast** (Camelyon16) [18], employed for metastasis detection in breast cancer, exemplifies a classic binary classification task in WSI analysis. It comprises 399 WSIs with or without metastasis, each accompanied by detailed pixel-level annotations.

**Lung** (TCGA-LU) employed for subtyping in non-small cell lung cancer, comprises a total of 937 WSIs, categorizing them into two subtypes: adenocarcinoma (LUAD) [19] and squamous cell carcinoma (LUSC) [20]. The cancerous region of 523 WSIs was entirely annotated at the pixel level by two experienced pathologists and four medical students.

**Ovarian** (UBC-OCEAN) [25] employed for subtyping in ovarian cancer, comprises a total of 513 WSIs, categorizing them into five subtypes: high-grade serous cancerous (HGSC), low-grade serous cancerous (LGSC), endometrioid cancerous (EC), clear cell carcinoma (CC), and mucinous cancerous (MC). Part of the cancerous, healthy, or necrotic regions of 152 WSIs were annotated, we combined the healthy and necrotic annotations to categorize them as normal tissue.

**RCC-3** (TCGA-RCC) includes a total of 660 WSIs, with three subtypes, clear cell RCC (CCRCC) [21], papillary RCC (PRCC) [22], and chromophobe RCC (CHRCC) [23]. The cancerous region of 338 WSIs was entirely annotated at the pixel level by two experienced pathologists and four medical students.

**RCC-4** (IH-RCC) collected from the First Affiliated Hospital of Xi’an Jiaotong University, with ethical approval (KYLLSL2021-420). It encompasses 563 WSIs from 168 patients across four RCC subtypes, CCRCC, PRCC, CHRCC, and Renal Oncocytoma (ROCY), with approximately 40 patients per subtype. The cancerous region of 138 WSIs was entirely annotated at the pixel level. **Gastric Endoscopy** (IH-ESD) collected from the First Affiliated Hospital of Xi’an Jiaotong University, with ethical approval (KYLLSL2022-333). It includes 99 patients with early gastric cancer Endoscopic Submucosal Dissection (ESD) specimens, a total of 286 WSIs. Each WSI was meticulously annotated at the pixel level, with three categories, *i*.*e*., tumor, inflammation, and normal tissue.

**Gastric** (TCGA-STAD) [26], comprising 339 Whole Slide Images (WSIs), was sourced from the TCGA Database. As the TCGA database does not provide detailed classification information for gastric adenocarcinoma, two pathologists with over a decade of experience classified all WSIs following the World Health Organization (WHO) histological classification system [45]. At the same time, they performed a detailed, patch-level annotation on 128 WSIs, identifying three tissue subtypes: highly differentiated (papillary and tubular), poorly differentiated, and mucinous.

**Prostate** (SICAPv2) [27], a publicly available prostate Gleason grading dataset, includes 153 WSIs labeled with categories G3, G4, G5, and normal tissue. This dataset provides tessellated patches with corresponding coordinates. Although the majority of patches come with annotations, a notable subset is devoid of labels. Two experienced pathologists provided supplementary annotations for these unlabelled patches.

Except for the already tessellated Prostate dataset, we process these WSIs into tiles of 2048×2048 pixels for 40x magnification and 1024×1024 pixels for 20x magnification without overlapping, thereby standardizing the input for consistent model training and evaluation. However, the Breast dataset presents a unique case. Given the variable sizes of tumor metastasis regions in this dataset, where some WSIs exhibit remarkably small tumor regions, we opt for a finer tessellation resolution of 512×512 pixels for the WSIs from this dataset. For the tessellated patches, we assign labels based on the predominant category within their representative regions, provided that pixel-level annotations are available. Additionally, some datasets (Breast, Gastric Endoscopy, and Prostate) include WSIs classified under the “Normal” category, indicating that all tessellated patches from these WSIs are normal (negative instance). Refer to **Supplementary Table 1** for statistics of these datasets.

### 4.4 Implementation details

Patch embeddings used for all methods are extracted from (the third residual block of the ResNet-50, which has been pre-trained on the ImageNet dataset. Where possible, configurations were aligned with the original implementations or their corresponding publications. It is, however, important to underline that the majority of these baselines are not intrinsically designed to accommodate multi-label classification tasks. Thus, modifications were introduced to these methods, with a particular emphasis on enhancing the attention aggregation mechanism, thereby extending their functionality to support multi-label classification. There is an exception, Trans-MIL is based on the self-attention mechanism and cannot be modified to multi-class attention. Therefore, in the experiment, TransMIL is unable to output patch-level predictions in multi-label datasets. Also, a weighted sampling technique is incorporated during the sample selection phase for all baselines, including the proposed SMMILe, to mitigate the issue of class imbalance. Furthermore, except for SMMILe, which possesses an instance refinement network capable of directly generating patch-level predictions, the derivation of patch-level predictions in representation-based attention MIL baselines, such as RAMIL, CLAM, DSMIL, TransMIL, and DTFD-MIL, relies on the raw attention scores. In the case of NIC and NIC-WSS, patch-level predictions are acquired from grad-CAM outputs, whereas for IAMIL, AddMIL, and the variants of SMMILe without integrating with the instance refinement network (Section 2.4), predictions are based on instance scores.

In the configuration of SMMILe, the kernel size of convolutional layer *cov*(·) is set to 1 × 1 for the Camelyon16 dataset, and 3 × 3 for other datasets. The super-patches are generated using the Simple Linear Iterative Clustering (SLIC) over-segmentation algorithm, as implemented in the scikit-image package. Considering both the high spatial homogeneity and the significant proportion of tumor areas in WSIs of multi-class classification datasets, the initial size of super-patches is set to 5×5 for multi-class datasets and 3×3 for others. Also, the integration of instance sampling with dropout is specifically utilized for multi-class classification datasets. The total number of sampling rounds *T* is set to 10, and the control parameters for the MRF constraint, *λ*_1_ and *λ*_2_, are fixed at 0.8 and 0.2, respectively, for all datasets.

The training of SMMILe is divided into two stages. In the first stage, the primary network of SMMILe is trained using the consistency loss ℒ_*cons*_ and the bag-level classification loss ℒ_*cls*_ for a maximum of 200 epochs. In the second stage, the number of linear layers *N* in the instance refinement network is set to 3, and the sample selection rate *θ* is established at 10 percent for each category. Both the primary and instance refinement networks are then trained using all loss functions, including ℒ_*cons*_, ℒ_*cls*_, ℒ_*ref*_, and ℒ_*mrf*_, for a maximum of 100 epochs. The Adam optimizer is used with a learning rate of 2*e*^*−*5^, and an early stopping strategy is implemented for both stages. For inference, the bag-level output score *P*_*c*_ for each category and the instance-level output scores 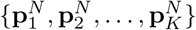 from the last linear layer *v*_*N*_ (·) are employed as the prediction results for the WSI and the corresponding patches, respectively. For all experiments, we used patient-level 5-fold cross-validation to estimate the predictive performance of each model. For each fold, four-fifths of the data were used to create a train-validation split (90%-10%), and the remaining fifth of the data was used as a test set. We use PyTorch for model implementation and the experiments are performed on a workstation with two NVIDIA 2080 Ti GPUs.

## Data Availability

All data produced in the present study are available upon reasonable request to the authors

## 5 Data availability

TCGA data (TCGA-LU, TCGA-RCC, and TCGA-STAD) including whole-slide images and diagnostic labels are available at https://portal.gdc.cancer.gov. Other publicly available datasets can be accessed in their corresponding data portals: Camelyon16 (https://camelyon16.grand-challenge.org/), UBC-OCEAN (https://www.kaggle.com/competitions/UBC-OCEAN), SICAPv2 (https://data.mendeley.com/datasets/9xxm58dvs3/1), The pixel-level annotations for TCGA-LU and TCGA-RCC are partially available at https://sites.google.com/view/aipath-dataset/home. The rest of the pixel-level annotations for TCGA-LU and TCGA-RCC, patch-level annotations, and fine-grained subtype labels for TCGA-STAD will be released upon publication. Two in-house datasets (IH-RCC and IH-ESD) were collected with IRB approval for the current study, and there are no plans to make them publicly available.

## 6 Code availability

The code for training SMMILe will be released upon publication. We have documented the details of model architecture and training, guaranteeing the reproducibility of SMMILe within the research community.

## 7 Acknowledgements

We acknowledge funding and support from Cancer Research UK and the Cancer Research UK Cambridge Centre [CTRQQR-2021-100012], The Mark Foundation for Cancer Research [RG95043], GE HealthCare, and the CRUK National Cancer Imaging Translational Accelerator (NCITA) [A27066]. Additional support was also provided by the National Institute of Health Research (NIHR) Cambridge Biomedical Research Centre [NIHR203312] and EPSRC Tier-2 capital grant [EP/P020259/1]. The funders had no role in study design, data collection and analysis, decision to publish, or preparation of the manuscript.

## Supplementary

### 1 Proofs

#### Proof of Theorem 1.

Revisiting *f* (·), typically designed as a linear projection involving two trainable vectors, denoted as **w** and **b**. The attention scores are generated by *g*(·) with softmax normalization over the instances, thereby ensuring that 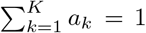 . Expanding Eq. (2), we derive:

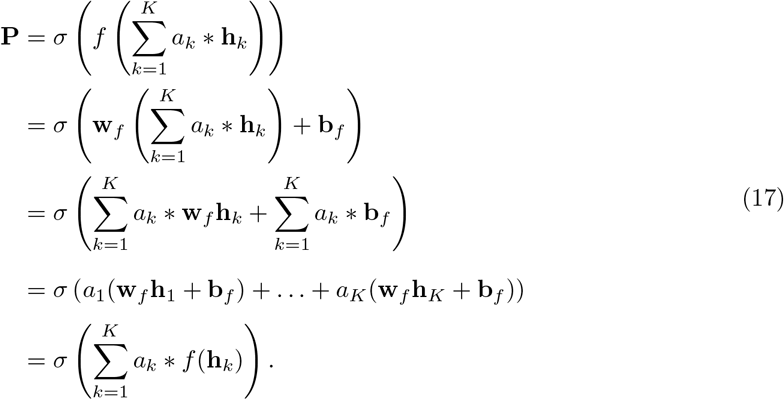

where the bag-level logit is aggregated as 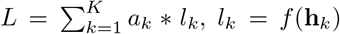. The resultant form represents a unique variant of IAMIL, which aggregates the raw outputs (logits) of *f* (·) followed by the activation function *σ*(·).

#### Proof of Theorem 2.

Utilizing the chain rule of differentiation, the partial derivative of 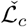 with respect to 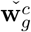 is formulated as:

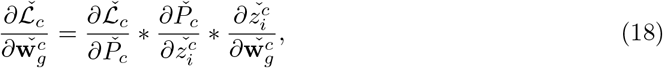

also, the partial derivative of 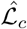 with respect to 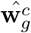 is described as:

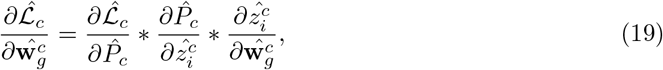

Moreover, the derivative of 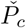 in relation to 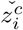 is computed as:

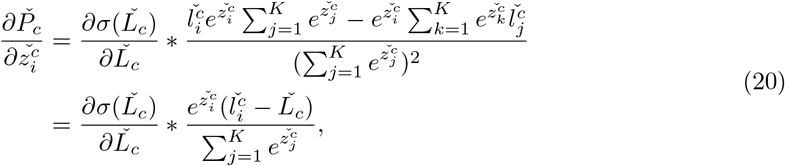

and the derivative of 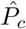 with respect to 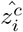 is calculated as:

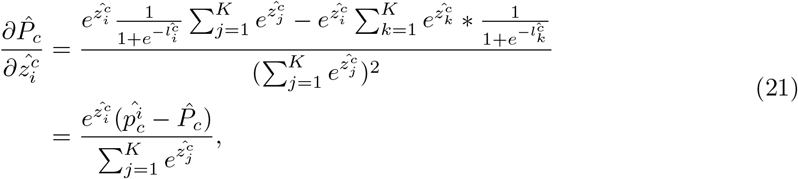

In light of Assumption 2, we deduce:

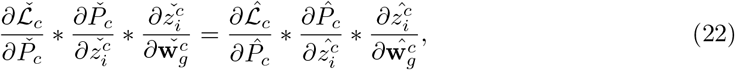

Where 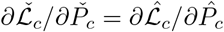. Furthermore, it can be readily demonstrated that 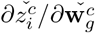 is eauiv-alent to 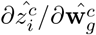 This equivalence arises because the values of these two partial derivatives are exclusively dependent on the architecture of the attention networks and the input vector **h**_*i*_ (with 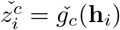 and 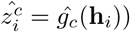, which remain consistent across RAMIL and IAMIL. Consequently, we can infer that:

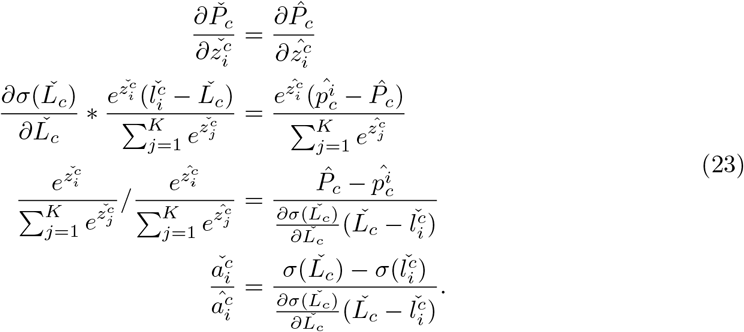

#### Proof of Theorem 3.

Firstly, The tangent line *t*(*l*) of *σ*(*l*) at *L*_*c*_ can be mathematically expressed as:

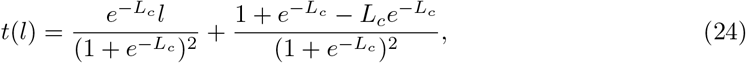

Given that the sigmoid function *σ*(*l*) has a single inflection point at *l* = 0, and asymptotic property towards positive and negative infinity, it can be inferred that the tangent line *t*(*l*) intersects with *σ*(*l*) at an additional point, denoted as (*L*_*int*_, *σ*(*L*_*int*_)), where *t*(*L*_*int*_) = *σ*(*L*_*int*_). Importantly, the product of *L*_*c*_ and *L*_*int*_ is negative, *i*.*e*., *L*_*c*_ · *L*_*int*_ *<* 0. The sole exception to this rule occurs when *L*_*c*_ = 0, in which case there is only one intersection point, *L*_*c*_ = *L*_*int*_ = 0. Moreover, it can be readily demonstrated using the geometric approach (**Suppplementary Fig**.

**1**) that if 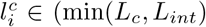, max(*L, L*)), then 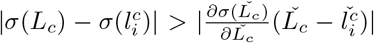, and vice versa.

**Supplementary Fig. 1:**
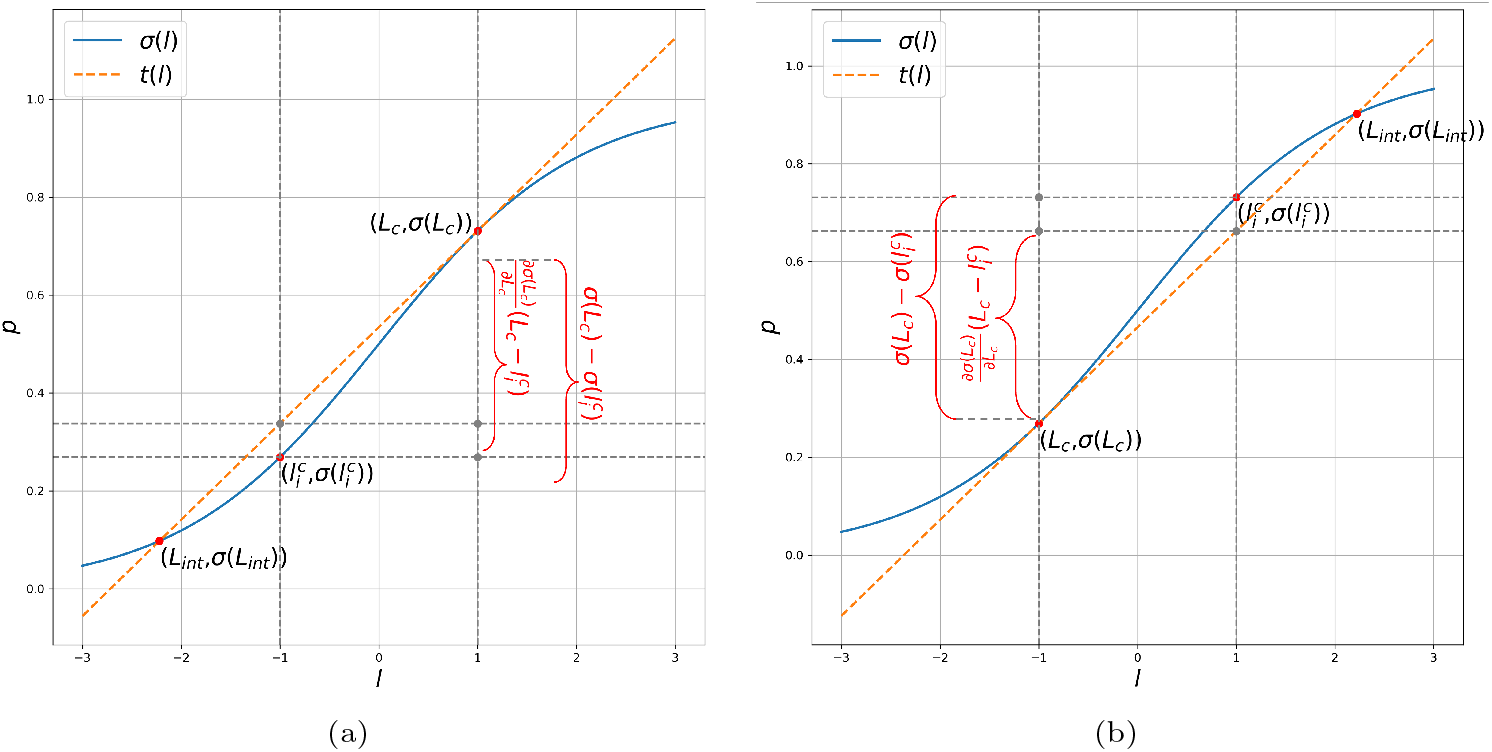
In conditions where *L*_*c*_ is (a) greater than 0, and (b) less than 0, the geometric proof regarding Theorem 3 is presented.

Secondly, the intersection point *L*_*int*_ can be expressed as:

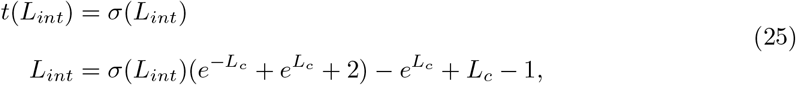

which is related to *L*_*c*_. However, it does not have an analytical solution. Alternatively, we analyze the properties of Eq. (25) and give a series of discrete solutions using Newton’s method. The first-order derivative of Eq. (25) with respect to *L*_*c*_ can be expressed as:

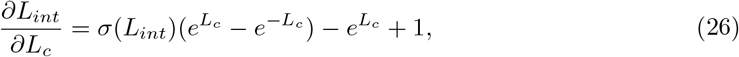

with Taylor expansion of 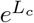 and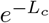, we have:

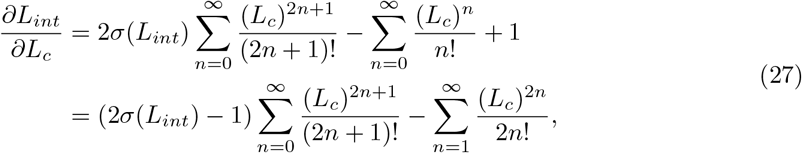

The second-order derivative of Eq. (25) with respect to *L*_*c*_ can be expressed as:

**Supplementary Fig. 2:**
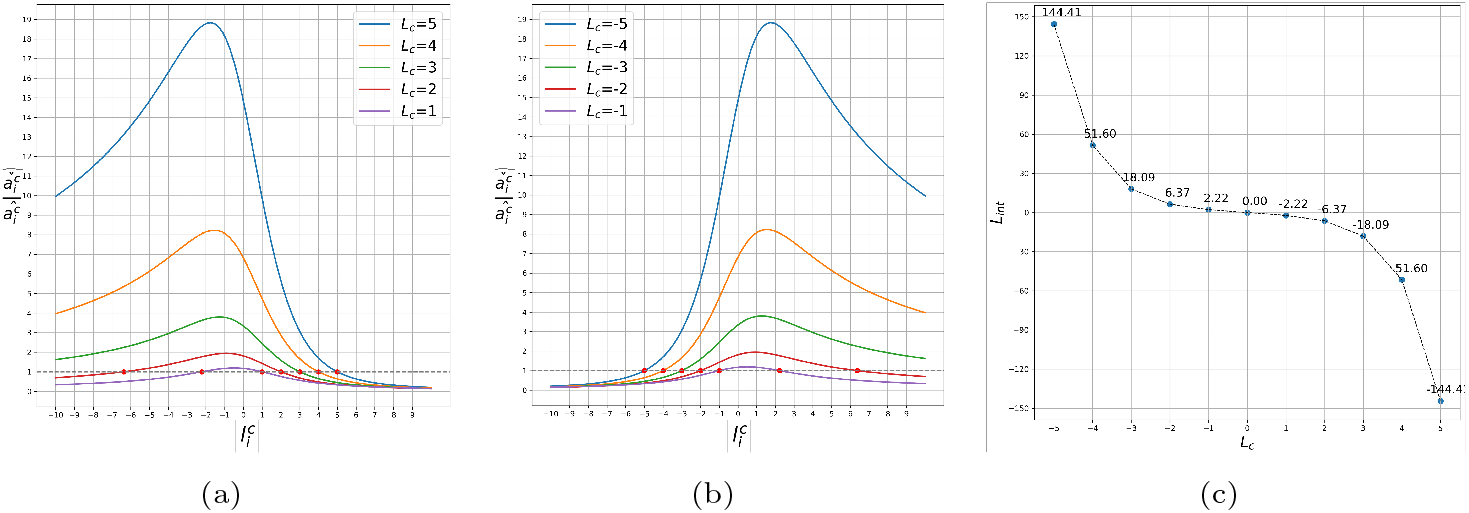
(a) Estimation results of *L*_*int*_ using Newton’s method with different *L*_*c*_. The ratio of 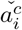 to 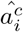 with different 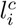 and *L*_*c*_, (b) when *L*_*c*_ *>* 0, (c) when *L*_*c*_ *<* 0.

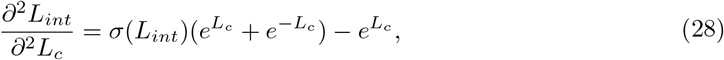

with Taylor expansion of 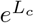 and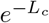, we have:

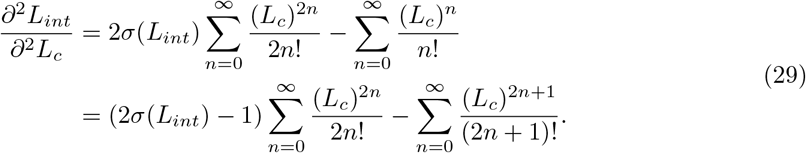

Since *L*_*c*_ · *L*_*int*_ *<* 0, if *L*_*c*_ *>* 0, it follows that (2*σ*(*L*_*int*_) − 1) *<* 0, indicating that *∂L*_*int*_*/∂L*_*c*_ *<* 0 and *∂*^2^*L*_*int*_*/∂*^2^*L*_*c*_ *<* 0. Similarly, if *L*_*c*_ *<* 0, the first-order derivative *∂L*_*int*_*/∂L*_*c*_ is still less than 0, but the second-order derivative *∂*^2^*L*_*int*_*/∂*^2^*L*_*c*_ *>* 0. If and only if *L*_*c*_ = 0, *∂L*_*int*_*/∂L*_*c*_ and *∂*^2^*L*_*int*_*/∂*^2^*L*_*c*_ are equal to 0. These properties of Eq. (25) indicate that (1) the distance between *L*_*c*_ and *L*_*int*_ increases as the absolute value of *L*_*c*_ increases, and (2) the rate of change for the absolute value of *L*_*int*_ also increases as the absolute value of *L*_*c*_ increases.

To elucidate the relationship between *L*_*c*_ and *L*_*int*_ more explicitly, we employed Newton’s method to solve Eq. (25). For *L*_*c*_ = {0, 1, 2, 3, 4, 5}, the corresponding *L*_*int*_ values are {0.00, −2.22, −6.37, −18.09, −51.60, −144.41}, respectively. For *L*_*c*_ *<* 0, the *L*_*int*_ values are the negatives of their respective positive counterparts (**Supplementary Fig. 2c**). The results reveal that as *L*_*c*_ varies, the distance between *L*_*c*_ and *L*_*int*_ changes at an almost exponential rate.

Furthermore, we calculated the ratio of 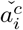 to 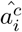 across different 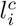 and *L*_*c*_ values (**Supplementary Fig. 2a,b**). It can be seen that when the absolute value of *L*_*c*_ is relatively large, the ratio of 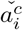 to 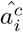 significantly exceeds 1 in most intervals, highlighting a substantial divergence between RAMIL and IAMIL in terms of attention scores allocated for low-discriminative and non-discriminative instances.

Considering the properties of the additional intersection point *L*_*int*_, along with the expected convergence of *σ*(*L*_*c*_) towards either 0 or 1, a relatively large magnitude of |*L*_*c*_| is often observed, typically exceeding 5 in most cases (**Supplementary Fig. 3a**). Consequently, this results in |*L*_*int*_| being greater than 100. It becomes evident that satisfying the condition 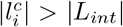 and 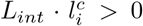is improbable. Moreover, for the majority of instances that meet the condition 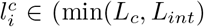, max(*L*_*c*_, *L*_*int*_)), the value of 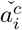 is expected to be several times greater than that of 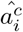.

Although these proofs have been proven using the sigmoid function, their applicability extends universally to the softmax function. Under the assumption that the raw outputs of other categories remain constant, and focusing solely on the relationship between the attention score 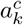 and the prediction score *P*_*c*_ for a single category, the softmax function can be effectively simplified to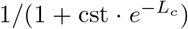, where “cst” represents a constant. Notably, this simplified representation of the softmax function exhibits properties analogous to those of the sigmoid function.

### 2 Hyperparameter Analysis

Here, we investigate the influences of two important hyperparameters for SMMILe on the Lung and Prostate datasets, (1) the initial size of super-patches for SLIC, from 2 × 2 to 6 × 6, and (2) The total number of instance sampling rounds, {1, 5, 10, 15}. We can see that both the WSI-level and patch-level classification performance of SMMILe is influenced by these two parameters (**Supplementary Fig. 5**). The impact on WSI-level classification performance is minimal, with variations generally within 1%, in contrast, patch-level classification performance experiences changes in the range of 3-4%. The number of instance sampling rounds has a negligible effect on the performance of SMMILe, except when the rounds are set to 1, where lower diversity leads to decreased performance, and settings of 5, 10, and 15 rounds yield similar results. On the other hand, the initial size of super-patches has a more significant impact on the performance of SMMILe. Both excessively large (6 × 6) and small (2 × 2) super-patches result in diminished performance, with the optimal parameters differing across datasets. For instance, the Lung dataset, characterized by larger WSI sizes and better spatial consistency, benefits more from larger sizes like 4 × 4 and 5 × 5, whereas in the Prostate dataset, where biopsy slide sizes are smaller, super-patches of 3 × 3 size perform better.

**Supplementary Table 1:** Settings for Synthetic Dataset Generation and Experimental Model Training.

| Setting | Value |
| --- | --- |
| Number of Classes | 2 |
| Embedding Dimension of Each Instance | 3 |
| Positive Distribution (Class 0) | Norm(-3.0,0.5);Norm(-2.0,0.5);Norm(3.0,0.5) |
| Positive Distribution (Class 1) | Norm(-2.0,0.5);Norm(-3.0,0.5);Norm(3.0,0.5) |
| Negative Distribution | Norm(1.0,0.5);Norm(-2.5,0.5);Norm(2.0,0.5) |
| Number of Bags per Class | 200 |
| Number of Instances per Bag | 1000 |
| The Ratio of Positive Instance per Bag | [0.1,1.0] |
| Attention Mechanism | Gated |
| Attention Network Feature Dimensions | 8, 4 |
| Dropout Rate | 0.25 |
| Optimizer | Adam |
| Learning Rate | 1e-3 |
| Weight Decay | 1e-5 |
| Stop Point (BCE loss) | 0.05 |

**Supplementary Fig. 3:**
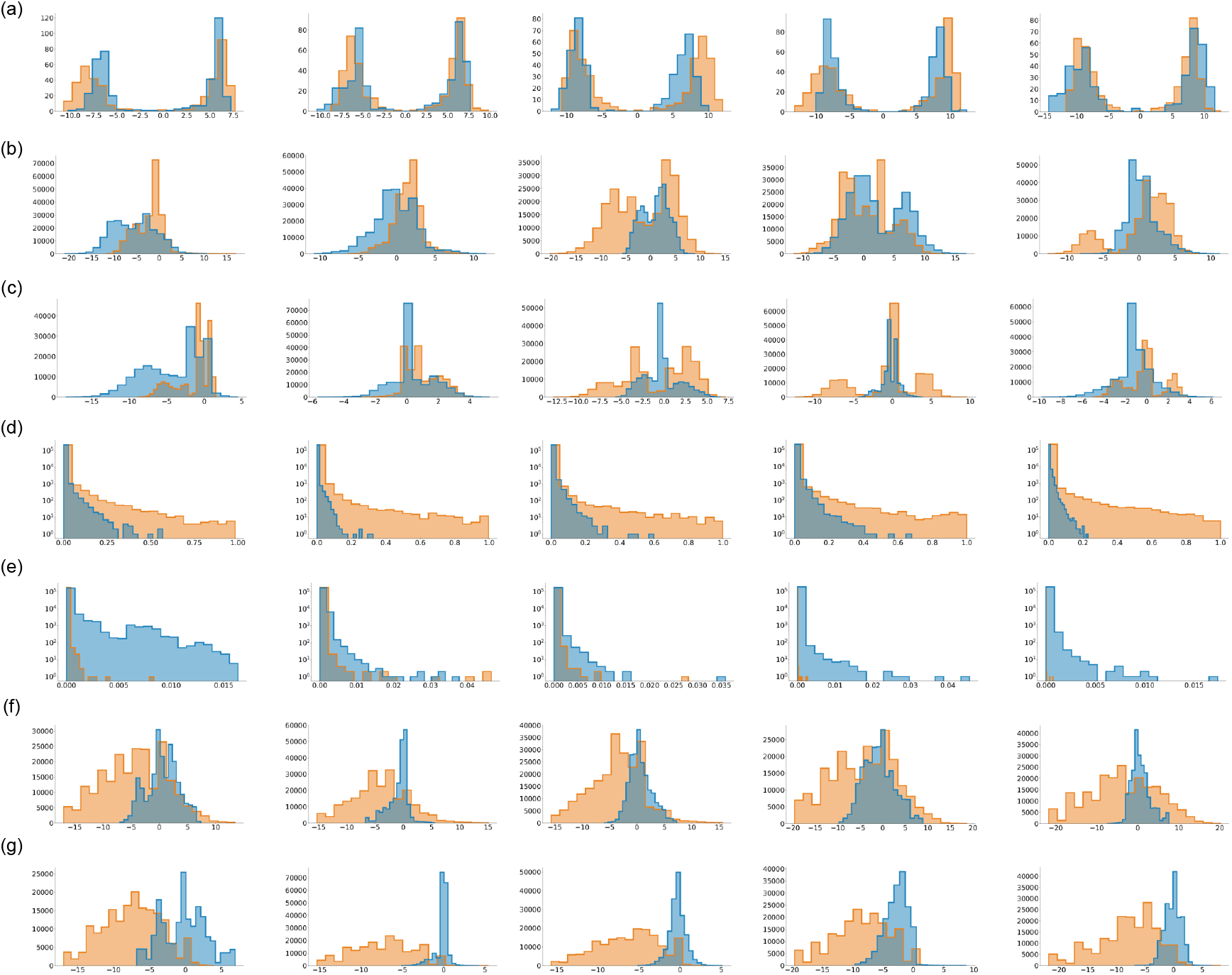
Histograms for five times synthetic experiments: (a) *L*_*c*_ for bags; (b) 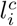, (d) 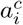, and (f) 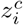 for positive instances; (c) 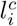, (e) 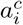, and (g) 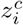 for negative instances. The y-axes of histograms for 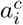 (d) and (e) are on a logarithmic scale.

**Supplementary Table 2:**
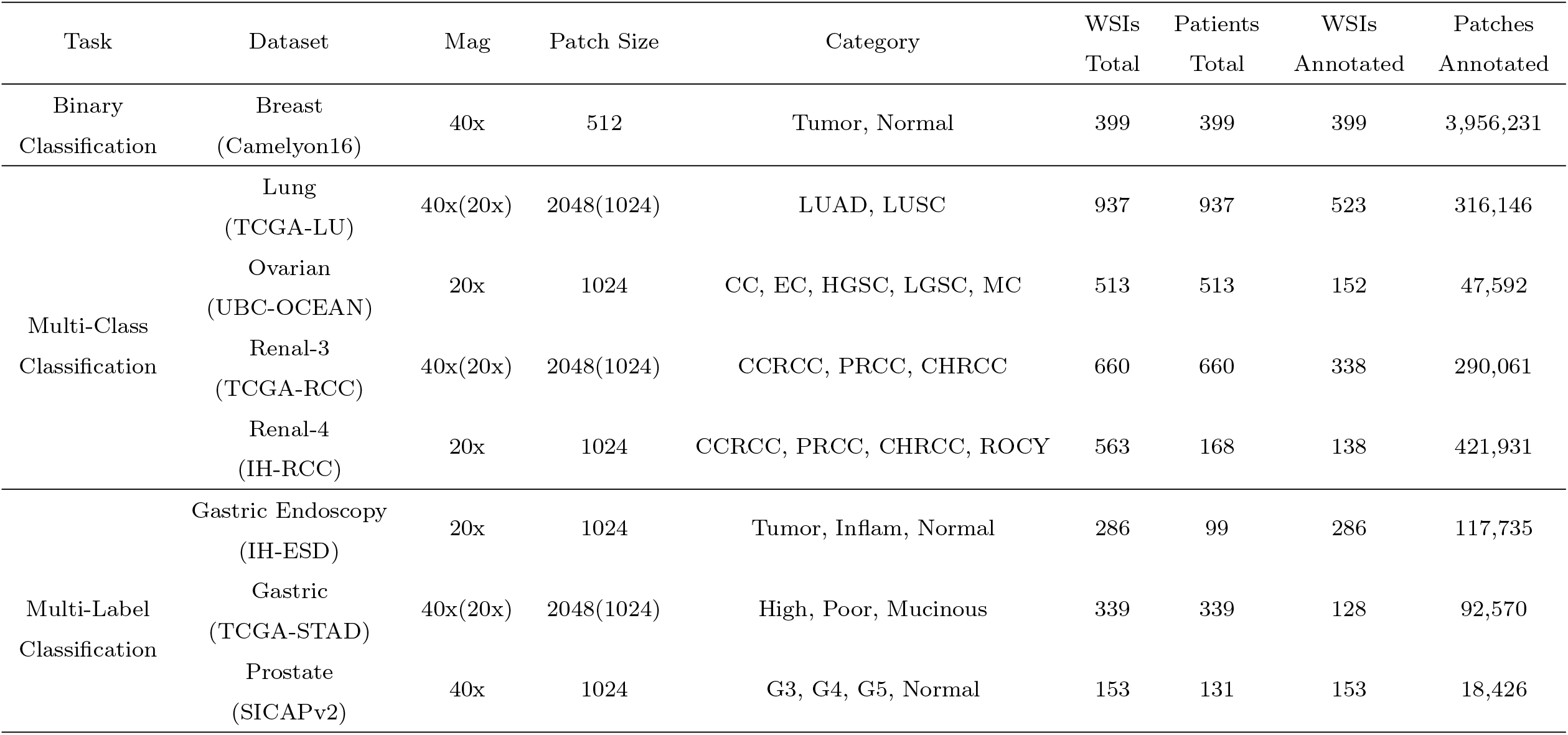
Statistics of the Datasets Used in Experiments, “WSIs Annotated” denotes WSIs with patch-level labels.

**Supplementary Table 3:** The comparison results (present in %) on Breast (Camelyon16). The best results are highlighted in boldface.

| Method | WSI Classification |  | Spatial Quantification |  |
| --- | --- | --- | --- | --- |
|  | Acc | AUC | Acc | AUC |
| RAMIL | 83.45±7.69 | 87.74±3.90 | 57.71±9.29 | 75.81±2.89 |
| IAMIL | 82.76±3.71 | 89.66±5.08 | 93.33±5.59 | 78.23±3.59 |
| CLAM | **83.96**±5.84 | 87.98±4.45 | 60.91±9.60 | 76.34±3.51 |
| DSMIL | 74.94±4.72 | 84.00±5.90 | 85.22±3.95 | 81.74±7.68 |
| TransMIL | 80.46±5.38 | 87.23±5.91 | 66.71±7.29 | 72.32±4.21 |
| DTFD-MIL | 83.21±3.24 | 90.05±4.61 | 67.54±9.60 | 79.79±3.51 |
| AddMIL | 60.41±3.19 | 77.81±4.21 | 53.68±3.58 | 58.59±5.56 |
| NIC | 42.85±6.39 | 43.82±3.50 | 65.74±5.35 | 65.47±4.90 |
| NIC-WSS | 41.87±5.26 | 46.79±4.52 | 66.42±4.48 | 59.45±6.49 |
| SMMILe | 82.43±2.75 | **90.51**±3.27 | **95.70**±1.61 | **89.28**±2.06 |

**Supplementary Table 4:** The comparison results (present in %) on Lung (TCGA-LU). The best results are highlighted in boldface.

| Method | WSI Classification |  | Spatial Quantification |  |
| --- | --- | --- | --- | --- |
|  | Acc | AUC | Acc | AUC |
| RAMIL | 82.85±3.54 | 90.44±1.89 | 50.10±3.13 | 57.42±3.44 |
| IAMIL | 80.81±3.06 | 88.55±2.05 | 46.47±3.37 | 58.86±2.97 |
| CLAM | 83.77±3.88 | 90.74±2.83 | 53.48±2.83 | 57.22±3.73 |
| DSMIL | 84.60±1.07 | 91.62±1.13 | 56.36±3.06 | 54.84±3.12 |
| TransMIL | 81.26±2.11 | 88.64±1.89 | 48.15±4.39 | 56.75±3.97 |
| DTFD-MIL | 81.01±0.66 | 89.63±1.59 | 58.33±3.63 | 61.79±2.34 |
| AddMIL | 78.74±2.16 | 83.79±2.94 | 42.22±3.86 | 49.11±3.49 |
| NIC | 48.15±0.85 | 62.17±3.07 | 57.13±5.02 | 50.87±3.14 |
| NIC-WSS | 64.78±9.63 | 76.11±7.83 | 63.88±2.82 | 63.73±0.99 |
| SMMILe | **87.68**±0.82 | **93.72**±1.01 | **73.35**±2.83 | **85.84**±0.93 |

**Supplementary Table 5:** The comparison results (present in %) on Ovarian (UBC-OCEAN). The best results are high-lighted in boldface.

| Method | WSI Classification |  | Spatial Quantification |  |
| --- | --- | --- | --- | --- |
|  | Acc | AUC | Acc | AUC |
| RAMIL | 75.08±5.15 | 91.60±2.72 | 75.46±8.85 | 86.59±2.48 |
| IAMIL | 66.85±9.69 | 89.58±4.30 | 13.28±1.94 | 81.10±10.51 |
| CLAM | 73.86±3.07 | 91.58±2.23 | 69.23±8.13 | 79.85±4.15 |
| DSMIL | **75.84**±5.86 | 92.54±2.35 | 57.30±7.87 | 73.22±3.70 |
| TransMIL | 65.20±7.75 | 87.89±2.95 | 19.44±6.44 | 46.44±3.28 |
| DTFD-MIL | 65.69±4.51 | 87.58±3.69 | 73.05±14.78 | 89.29±5.61 |
| AddMIL | 69.99±3.82 | 89.80±2.27 | 12.46±2.31 | 61.67±7.95 |
| NIC | 45.41±6.27 | 80.64±6.78 | 53.87±2.26 | 43.95±9.28 |
| NIC-WSS | 51.72±4.05 | 83.59±3.34 | 57.56±1.12 | 42.23±5.95 |
| SMMILe | 75.64±4.59 | **93.21**±1.07 | **91.99**±0.50 | **92.58**±2.29 |

**Supplementary Table 6:** The comparison results (present in %) on Renal-3 (TCGA-RCC). The best results are highlighted in boldface.

| Method | WSI Classification |  | Spatial Quantification |  |
| --- | --- | --- | --- | --- |
|  | Acc | AUC | Acc | AUC |
| RAMIL | 90.63±3.08 | 97.52±1.36 | 49.59±2.56 | 50.75±2.34 |
| IAMIL | 90.01±1.65 | 97.31±0.69 | 43.88±4.16 | 62.04±2.72 |
| CLAM | 92.14±3.18 | 98.28±1.06 | 53.06±2.05 | 55.97±1.97 |
| DSMIL | 89.57±2.96 | 97.31±1.05 | 54.21±3.05 | 58.25±3.09 |
| TransMIL | 89.85±2.39 | 97.49±0.70 | 53.25±3.50 | 47.11±3.76 |
| DTFD-MIL | 89.48±4.37 | 96.71±1.90 | 59.80±3.81 | 56.47±3.46 |
| AddMIL | 88.45±3.32 | 96.95±1.56 | 43.49±4.47 | 47.85±5.29 |
| NIC | 85.15±3.02 | 94.47±4.62 | 50.69±7.24 | 48.97±4.89 |
| NIC-WSS | 86.97±1.73 | 97.07±0.89 | 64.63±6.12 | 68.57±8.26 |
| SMMILe | **94.63**±2.56 | **99.00**±0.63 | **78.77**±4.34 | **90.89**±4.39 |

**Supplementary Table 7:** The comparison results (present in %) on Renal-4 (IH-RCC). The best results are highlighted in boldface.

| Method | WSI Classification |  | Spatial Quantification |  |
| --- | --- | --- | --- | --- |
|  | Acc | AUC | Acc | AUC |
| RAMIL | 77.55±3.63 | 90.75±3.40 | 44.29±2.46 | 53.55±3.52 |
| IAMIL | 81.63±1.97 | 92.67±1.73 | 34.95±2.70 | 59.43±0.78 |
| CLAM | 80.38±3.19 | 92.33±2.70 | 59.04±2.43 | 60.14±2.05 |
| DSMIL | 82.70±2.12 | 93.28±2.48 | 48.93±3.49 | 51.31±4.64 |
| TransMIL | 81.69±2.90 | 92.91±1.87 | 42.46±5.26 | 48.96±3.64 |
| DTFD-MIL | 79.70±3.12 | 91.08±2.59 | 52.27±4.83 | 56.03±2.43 |
| AddMIL | 73.33±6.50 | 86.18±3.06 | 47.69±2.29 | 50.70±4.16 |
| NIC | 68.51±5.47 | 83.86±3.14 | 52.25±3.22 | 50.20±1.42 |
| NIC-WSS | 73.10±2.61 | 88.71±1.98 | 63.13±0.68 | 57.09±2.59 |
| SMMILe | **87.17**±1.46 | **94.96**±2.26 | **80.54**±1.76 | **91.07**±0.89 |

**Supplementary Table 8:** The comparison results (present in %) on Gastric Endoscopy (IH-ESD). The best results are highlighted in boldface.

| Method | WSI Classification |  | Spatial Quantification |  |
| --- | --- | --- | --- | --- |
|  | Acc | AUC | Acc | AUC |
| RAMIL | 88.29±3.17 | 86.82±6.40 | 58.66±1.14 | 63.21±2.04 |
| IAMIL | 87.12±2.99 | 86.77±6.23 | 55.38±1.07 | 67.08±8.38 |
| CLAM | 88.46±3.11 | 88.20±3.72 | 59.10±2.21 | 63.82±3.12 |
| DSMIL | 86.86±2.78 | 86.10±3.40 | 59.21±3.72 | 65.44±2.22 |
| TransMIL | 88.54±3.77 | 86.18±6.51 | - | - |
| DTFD-MIL | 83.78±0.47 | 81.71±6.84 | 54.44±2.86 | 62.70±2.14 |
| AddMIL | 86.12±2.19 | 85.25±3.92 | 46.22±6.22 | 50.13±3.42 |
| NIC | 81.65±2.72 | 77.09±3.56 | 48.81±2.85 | 51.68±3.24 |
| NIC-WSS | 79.03±2.65 | 77.43±8.38 | 48.25±1.09 | 51.99±2.74 |
| SMMILe | **90.36**±3.39 | **92.97**±3.14 | **61.86**±1.61 | **76.53**±2.79 |

**Supplementary Table 9:** The comparison results (present in %) on Gastric (TCGA-STAD). The best results are highlighted in boldface.

| Method | WSI Classification |  | Spatial Quantification |  |
| --- | --- | --- | --- | --- |
|  | Acc | AUC | Acc | AUC |
| RAMIL | 81.35±2.44 | 82.03±3.96 | 68.88±4.30 | 76.56±2.77 |
| IAMIL | 80.24±3.87 | 81.55±4.30 | 59.48±5.22 | 75.75±4.87 |
| CLAM | 81.83±3.21 | **83.51**±4.69 | 68.91±2.01 | 76.53±1.74 |
| DSMIL | 75.39±1.66 | 80.94±4.39 | 71.02±6.80 | 76.05±6.51 |
| TransMIL | 75.41±3.80 | 81.38±6.38 | - | - |
| DTFD-MIL | 73.90±1.90 | 77.49±3.95 | 50.88±4.65 | 49.78±3.30 |
| AddMIL | 74.01±1.04 | 79.60±3.66 | 36.08±3.71 | 42.56±6.23 |
| NIC | 76.86±3.86 | 76.11±4.36 | 60.35±3.15 | 62.36±5.63 |
| NIC-WSS | 68.38±5.17 | 69.63±7.00 | 52.52±7.17 | 56.82±3.61 |
| SMMILe | **82.54**±1.97 | 83.31±3.37 | **75.16**±1.64 | **89.70**±2.55 |

**Supplementary Table 10:** The comparison results (present in %) on Prostate (SICAPv2). The best results are highlighted in boldface.

| Method | WSI Classification |  | Spatial Quantification |  |
| --- | --- | --- | --- | --- |
|  | Acc | AUC | Acc | AUC |
| RAMIL | 83.29±3.65 | 88.68±3.00 | 65.45±1.67 | 66.65±1.46 |
| IAMIL | 84.40±4.38 | 89.44±2.60 | 62.31±2.76 | 71.52±1.15 |
| CLAM | 83.92±3.71 | 89.00±3.26 | 62.31±2.76 | 66.24±2.37 |
| DSMIL | 72.33±3.60 | 87.61±3.52 | 66.41±3.72 | 72.26±2.22 |
| TransMIL | 72.76±3.98 | 86.66±3.04 | - | - |
| DTFD-MIL | 70.18±2.95 | 82.96±4.07 | 60.73±2.99 | 56.29±4.48 |
| AddMIL | 70.61±3.41 | 85.01±4.82 | 52.70±3.51 | 52.98±2.48 |
| NIC | 79.44±3.27 | 79.72±5.78 | 63.03±2.07 | 67.43±2.44 |
| NIC-WSS | 67.81±6.10 | 70.59±5.36 | 52.32±1.04 | 57.94±1.50 |
| SMMILe | **87.32**±3.50 | **90.75**±3.56 | **72.38**±3.55 | **89.49**±2.66 |

**Supplementary Fig. 4:**
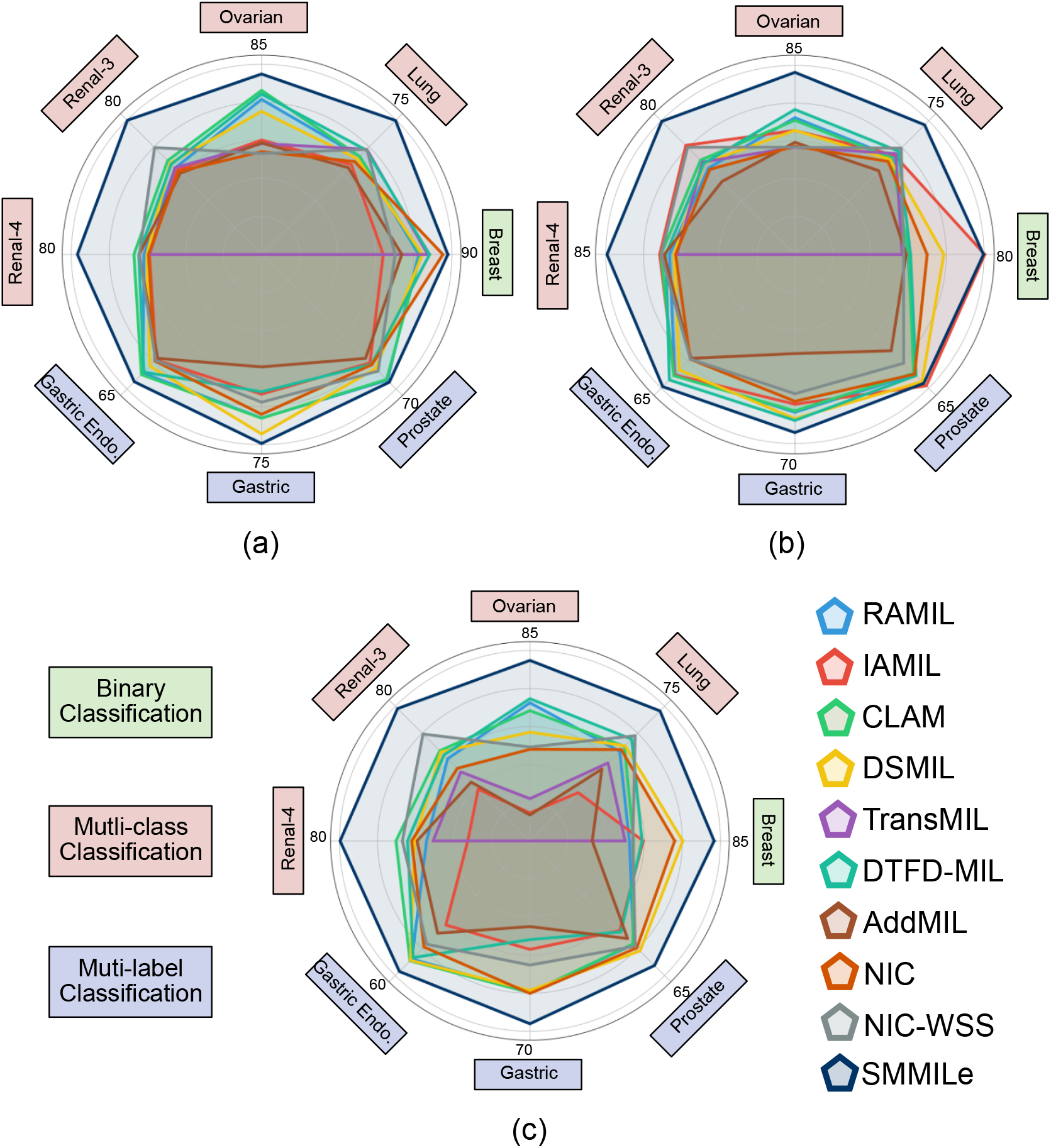
Spatial quantification results (present in %) for different methods across diverse datasets: (a) macro precision, (b) macro recall, (c) macro F1-score.

**Supplementary Fig. 5:**
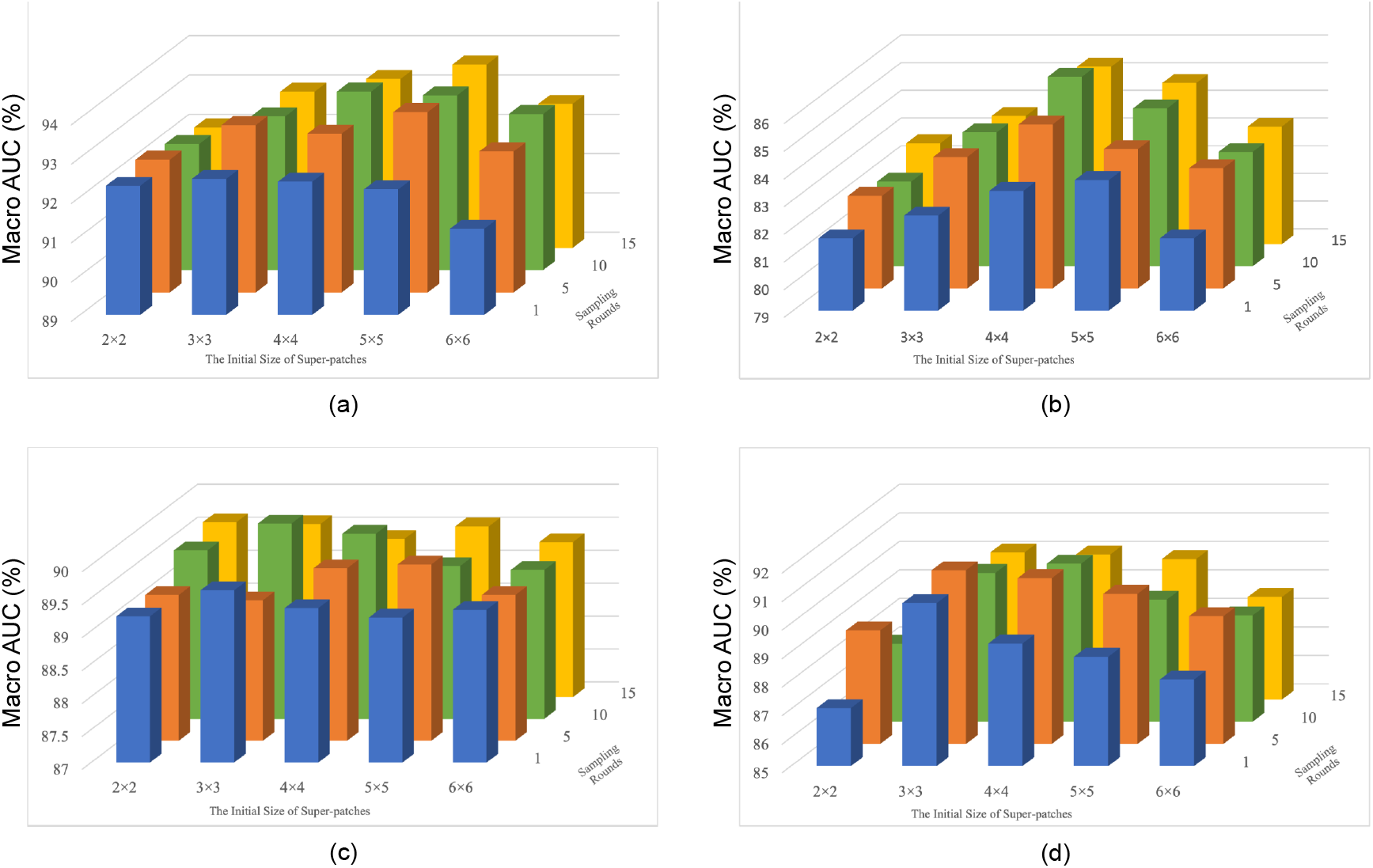
The WSI classification and spatial quantification results with different superpatch initial sizes and instance sampling rounds on (a,b) Lung (TCGA-LU) and (c,d) Prostate (SICAPv2) datasets, respectively.

**Supplementary Fig. 6:**
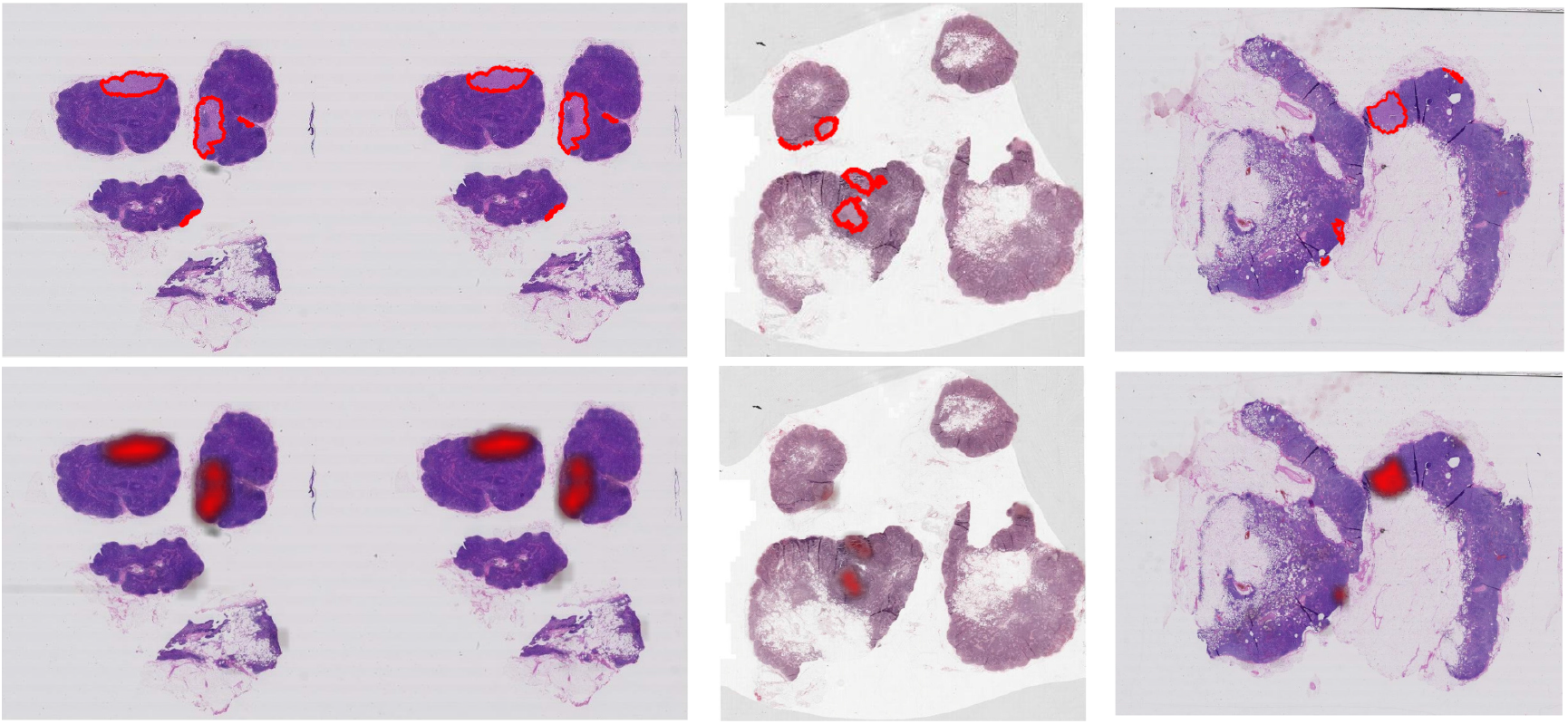
Visualization of spatial quantification results of cases from Breast (Camelyon16). GT in the first row.

**Supplementary Fig. 7:**
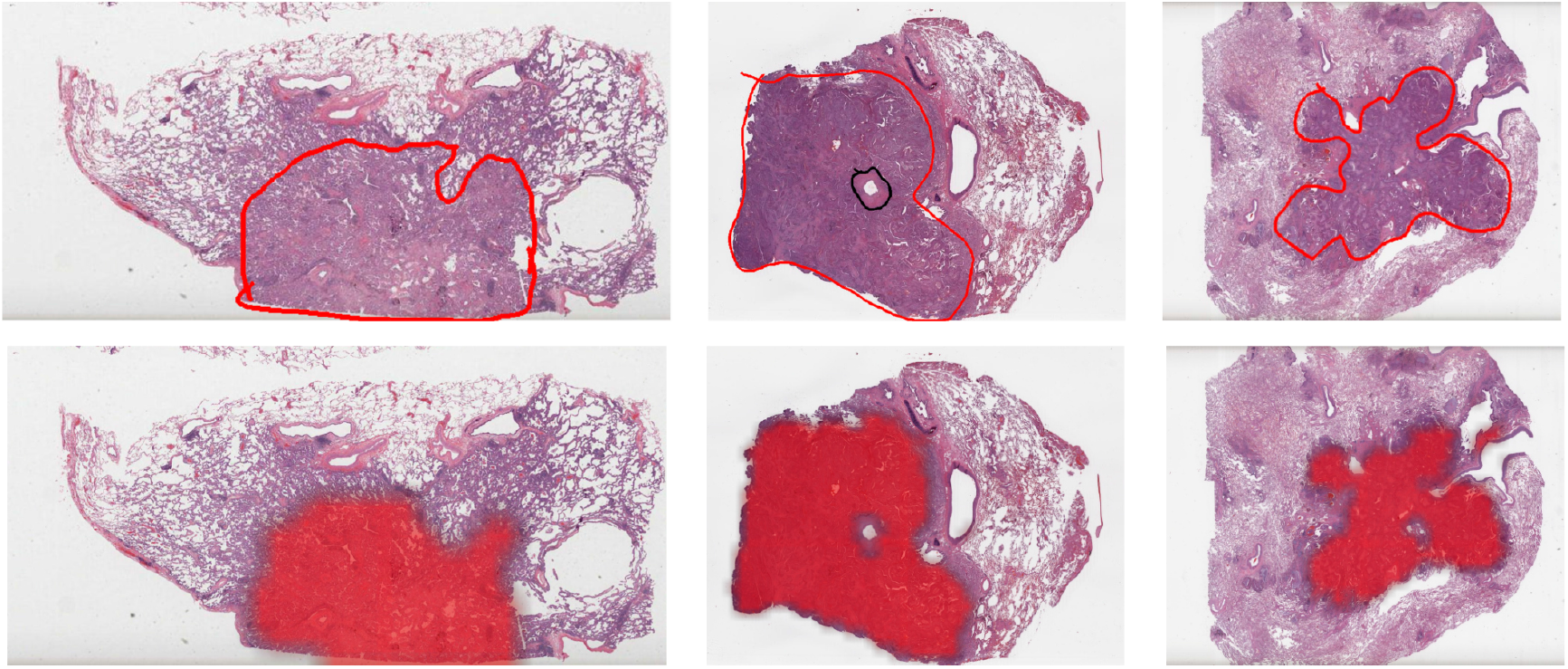
Visualization of spatial quantification results of cases from Lung (TCGA-LU). GT in the first row.

**Supplementary Fig. 8:**
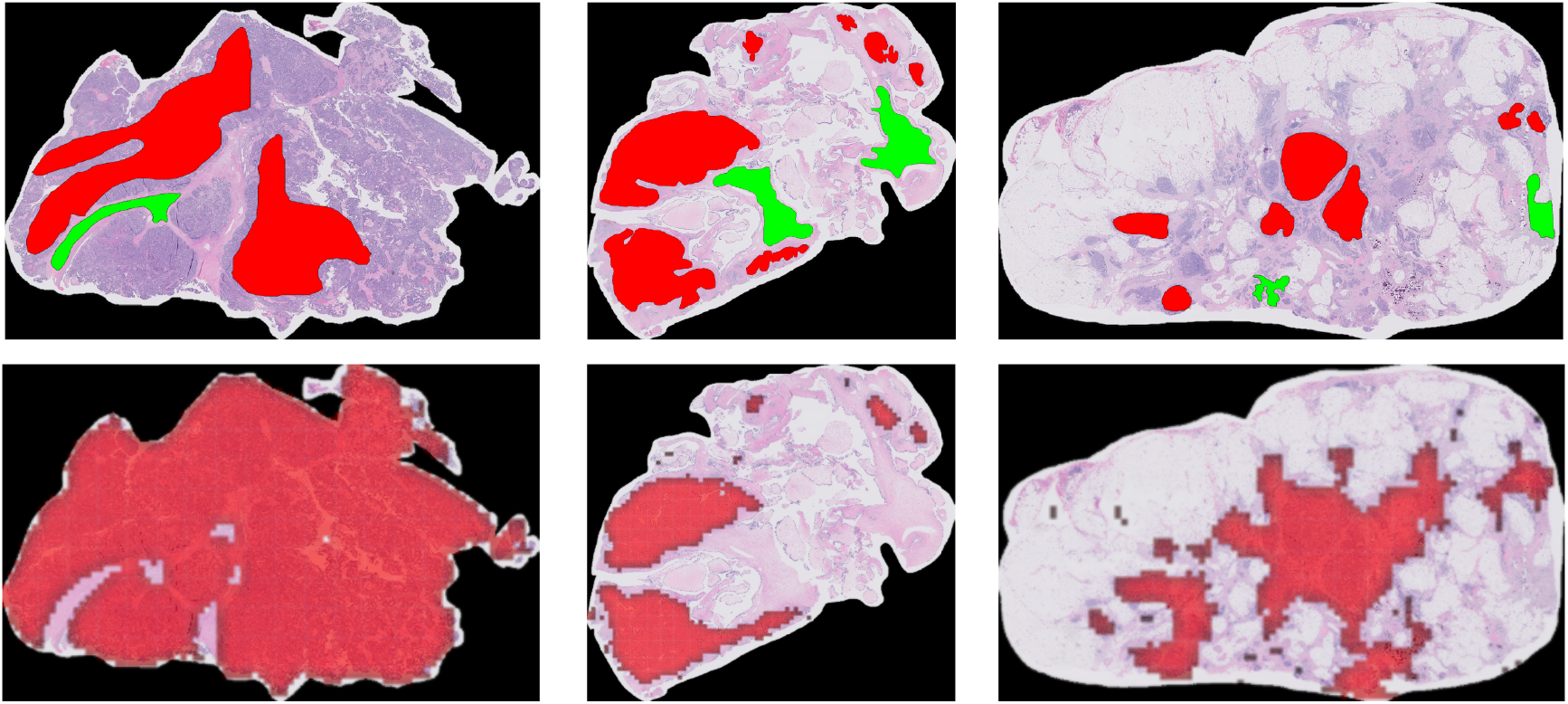
Visualization of spatial quantification results of cases from Ovarian (UBC-OCEAN). GT in the first row.

**Supplementary Fig. 9:**
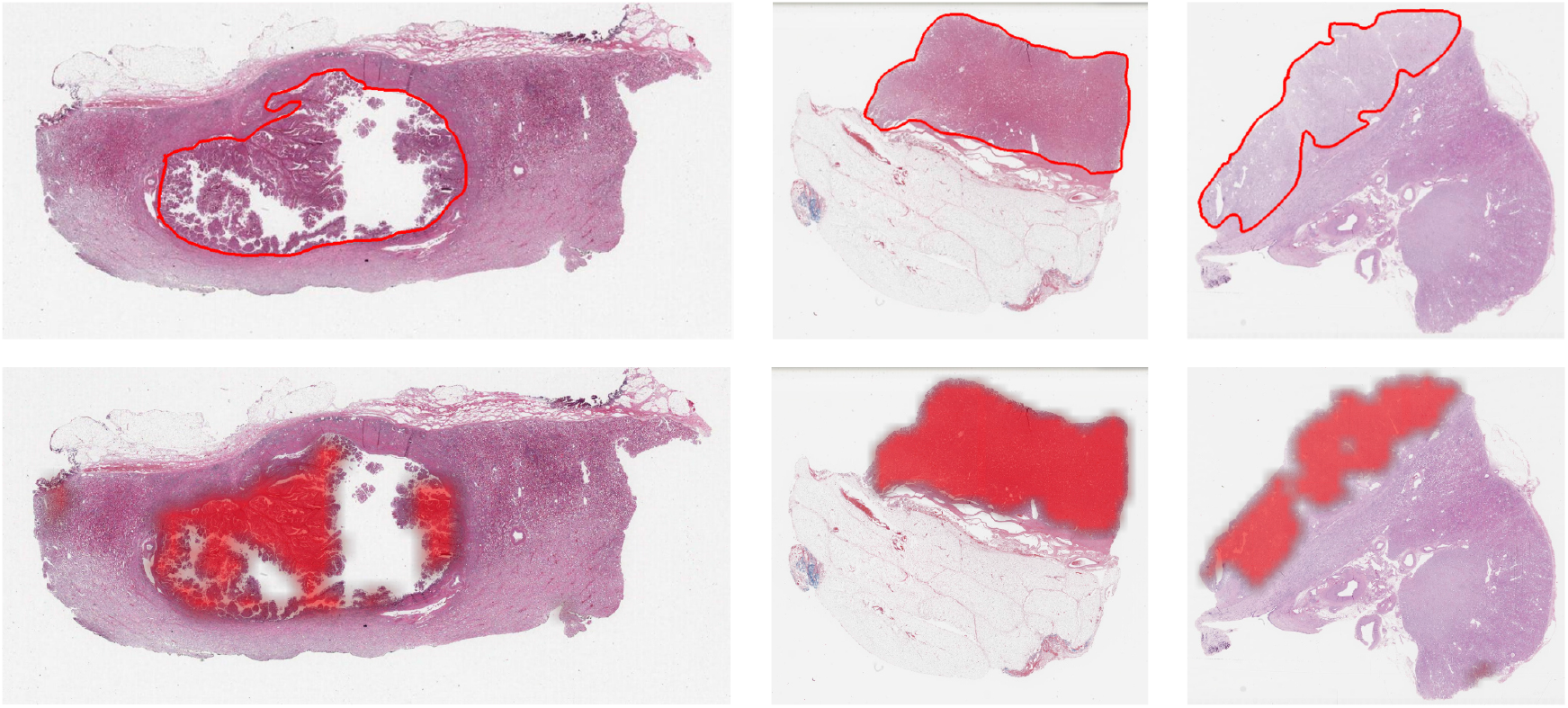
Visualization of spatial quantification results of cases from Renal-3 (TCGA-RCC). GT in the first row.

**Supplementary Fig. 10:**
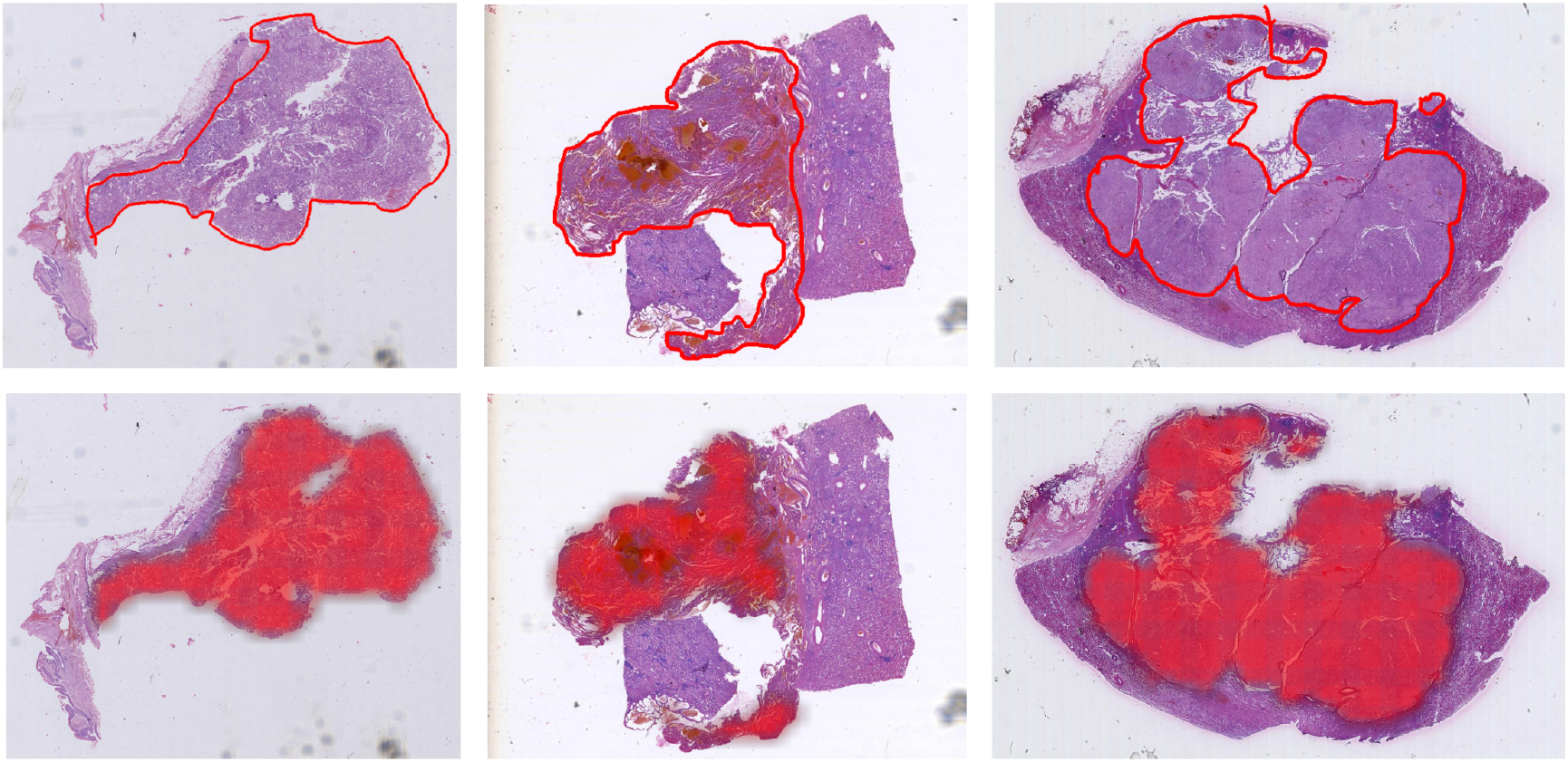
Visualization of spatial quantification results of cases from Renal-4 (IH-RCC). The annotated green color represents normal tissue. GT in the first row.

**Supplementary Fig. 11:**
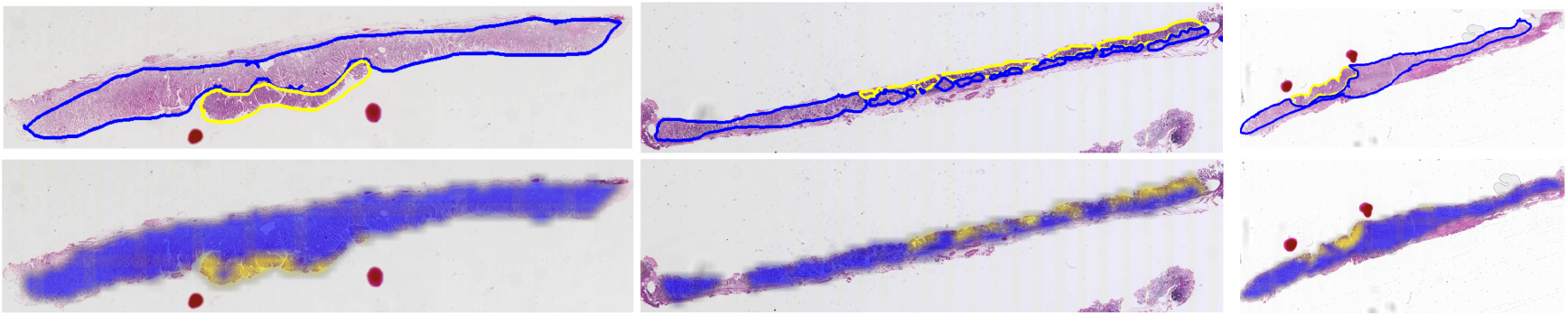
Visualization of spatial quantification results of cases from Gastric Endoscopy (IH-ESD). The annotated yellow and blue colors represent tumor and inflammation, respectively. GT in the first row.

**Supplementary Fig. 12:**
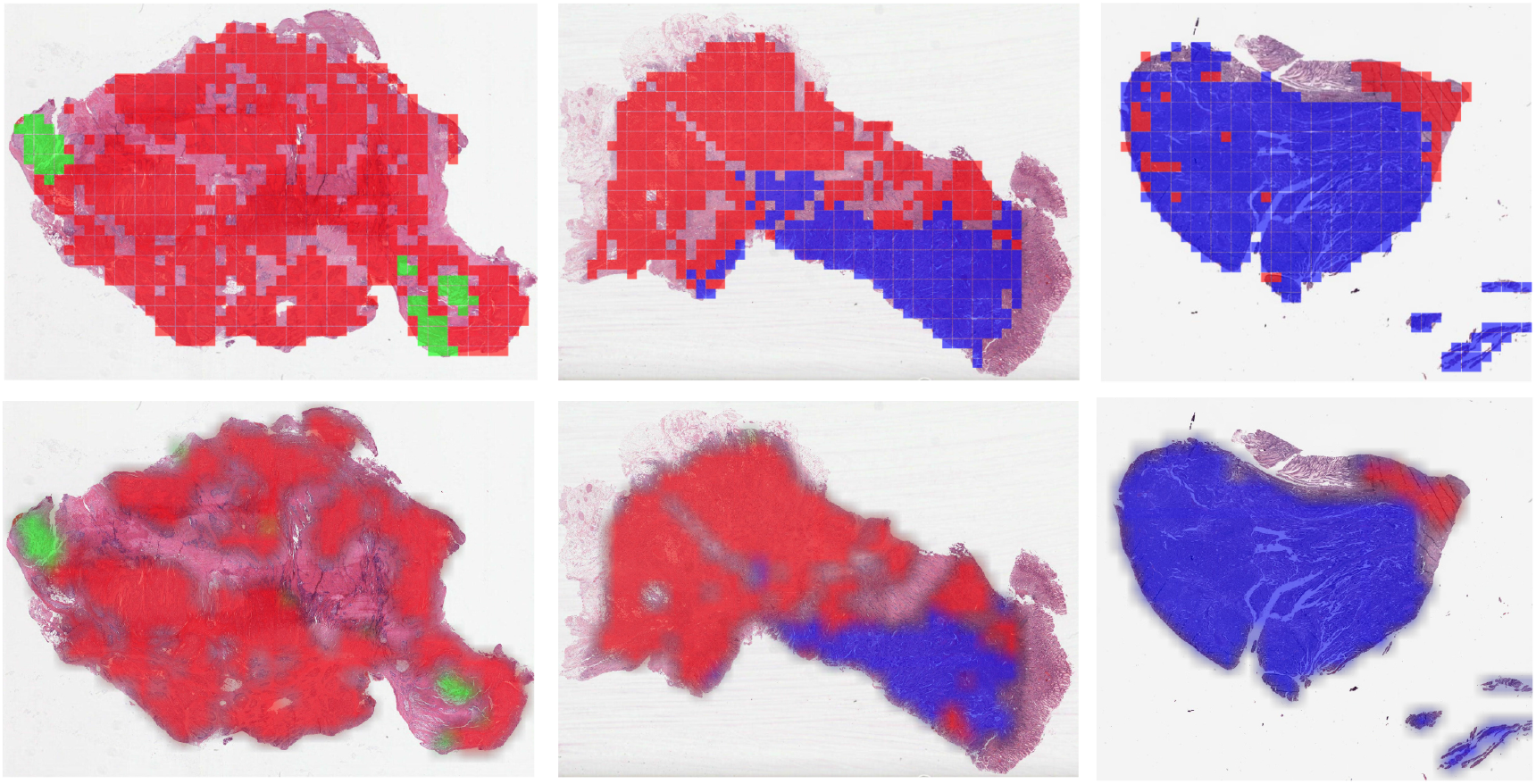
Visualization of spatial quantification results of cases from Gastric (TCGA-STAD). The annotated red, blue, and green colors represent highly-differentiate, poorly-differentiate, and mucinous, respectively. GT in the first row.

**Supplementary Fig. 13:**
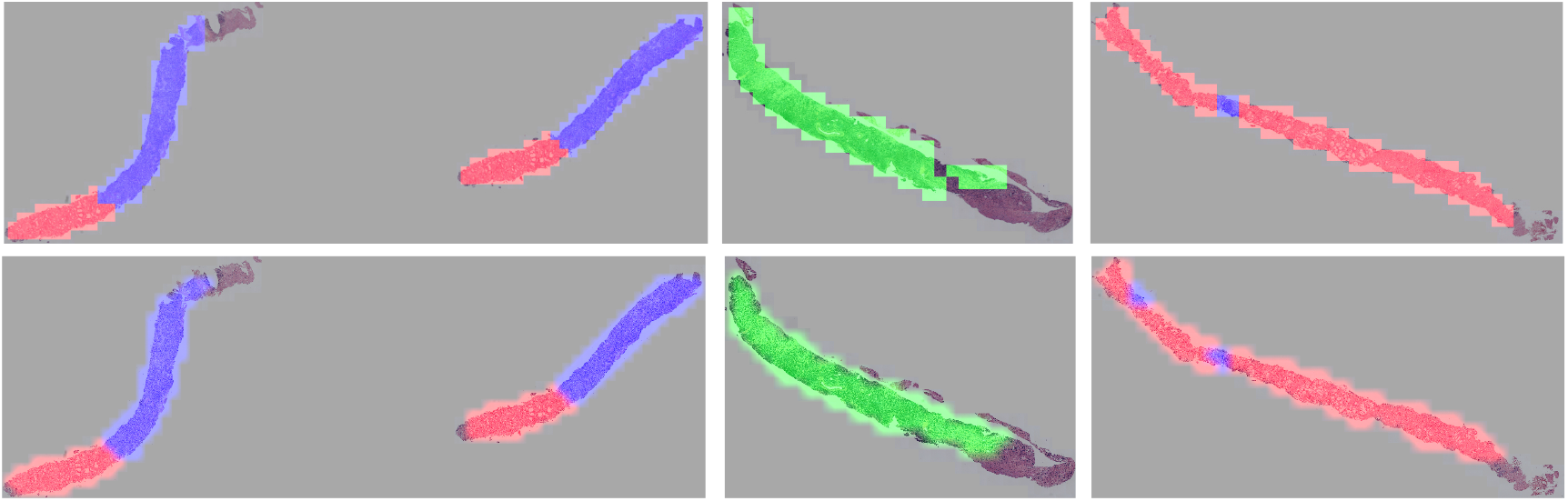
Visualization of spatial quantification results of cases from Prostate (SICAPv2). The annotated red, blue, and green colors represent G3, G4, and G5 Gleason grading, respectively. GT in the first row.

